# Plasma-based organ-specific aging and mortality models unveil diseases as accelerated aging of organismal systems

**DOI:** 10.1101/2024.04.08.24305469

**Authors:** Ludger J.E. Goeminne, Alec Eames, Alexander Tyshkovskiy, M. Austin Argentieri, Kejun Ying, Mahdi Moqri, Vadim N. Gladyshev

**Author notes:** Correspondence should be addressed to V.N.G.

## Abstract

Aging is a complex process manifesting at the molecular, cell, organ and organismal levels. It leads to functional decline, disease and ultimately death, but the relationship between these fundamental biomedical features remains elusive. By applying machine learning to plasma proteome data of over fifty thousand human subjects in the UK Biobank and other cohorts, we report organ-specific and conventional aging models trained on chronological age, mortality and longitudinal proteome data. We show how these tools predict organ/systems-specific disease through numerous phenotypes. We find that men are biologically older and age faster than women, that accelerated aging of organs leads to diseases in these organs, and that specific diets, lifestyles, professions and medications are associated with accelerated and decelerated aging of specific organs and systems. Altogether, our analyses reveal that age-related chronic diseases epitomize accelerated organ- and system-specific aging, modifiable through environmental factors, advocating for both universal whole-organism and personalized organ/system-specific anti-aging interventions.

## Introduction

Aging is a highly complex process that manifests across multiple scales, from molecular changes within cells to functional decline at the tissue, organ, and organismal levels, resulting in disease and increased mortality risk (Gladyshev et al., 2021). Understanding the mechanisms underlying this multifaceted process is crucial for developing interventions that promote healthspan and longevity. However, our current knowledge of how aging mechanisms across various levels interconnect and contribute to the development of age-related diseases remains limited.

Molecular aging clocks, initially developed based on DNA methylation patterns (Horvath, 2013), have been extended to other omics modalities, such as the transcriptome (Peters et al., 2015) and proteome (Lehallier et al., 2019). These aging clocks estimate an individual’s biological age, which has been defined as “an individual’s age defined by the level of age-dependent biological changes, such as molecular and cellular damage accumulation”. It may practically be “summarized as a number (in units of time) matching the chronological age where the average person in a reference population shares the individual’s level of age-dependent biological changes” (Moqri et al., 2023). As such, biological age can deviate from chronological age and this age deviation can serve as a biomarker for age-related diseases and mortality risk (Moqri et al., 2023). Epigenetic clocks trained on different tissues and organs can produce strikingly divergent outcomes, implying different organs age at distinct rates (Prattichizzo et al., 2024).

Recently, Oh et al. (2023) provided an excellent proof-of-concept that individual organs can age differently across human populations and showed that accelerated organ-specific aging can be captured by analyzing the plasma proteome. This study utilized several cohorts encompassing 5,676 individuals, but larger and more extensively phenotyped population-based cohorts, such as the UK Biobank, provide an unprecedented opportunity to uncover omics-based organ-specific signatures of aging and investigate the effects of lifestyle, diet and therapeutic interventions on aging-related health outcomes (Sudlow et al., 2015). They also enable the development of robust proteome-based biomarkers to assess biological age (Argentieri et al., 2023). The Olink Explore 3072 platform, a powerful high-throughput affinity-based proteomics platform that measures protein levels of ∼3,000 proteins simultaneously, has recently been applied to the plasma of UK Biobank participants (Eldjarn et al., 2023).

In this study, we leveraged plasma proteomics data from over 53,000 UK Biobank participants to develop organ-specific aging models. By annotating protein abundance patterns with organ-specific annotations from the Genotype-Tissue Expression (GTEx) project (Lonsdale et al., 2013), we identified proteomic signatures in the plasma that reflect differential aging rates of various organs and trained biomarkers based on chronological age and mortality. Their application to various phenotypes offers numerous insights into the human aging process at the organ level and suggests that chronic diseases generally manifest as accelerated organ- and system-specific aging, modifiable through lifestyle interventions, such as diets and medications.

## Main

### Plasma proteome supports robust chronological aging models

We randomly split the UK Biobank participants into a training set (42,412 UK Biobank participants, 80%) and a test dataset (10,603 UK Biobank participants, 20%) and trained an elastic net model on 2,920 plasma proteins in the training set based on the participants’ chronological age. Model coefficients for this 1^st^-generation model were non-zero for 2,246 of these proteins (Supp. Table 1). The age predicted by our model has a strong positive correlation with chronological age, both in the training (Fig. 1a, correlation coefficient r = 0.94, mean absolute error = 0.78 years) and test datasets (Fig. 1b, correlation coefficient r = 0.93, mean absolute error = 0.78 years). These characteristics are on par with gold standard epigenetic aging models.

**Figure 1.**
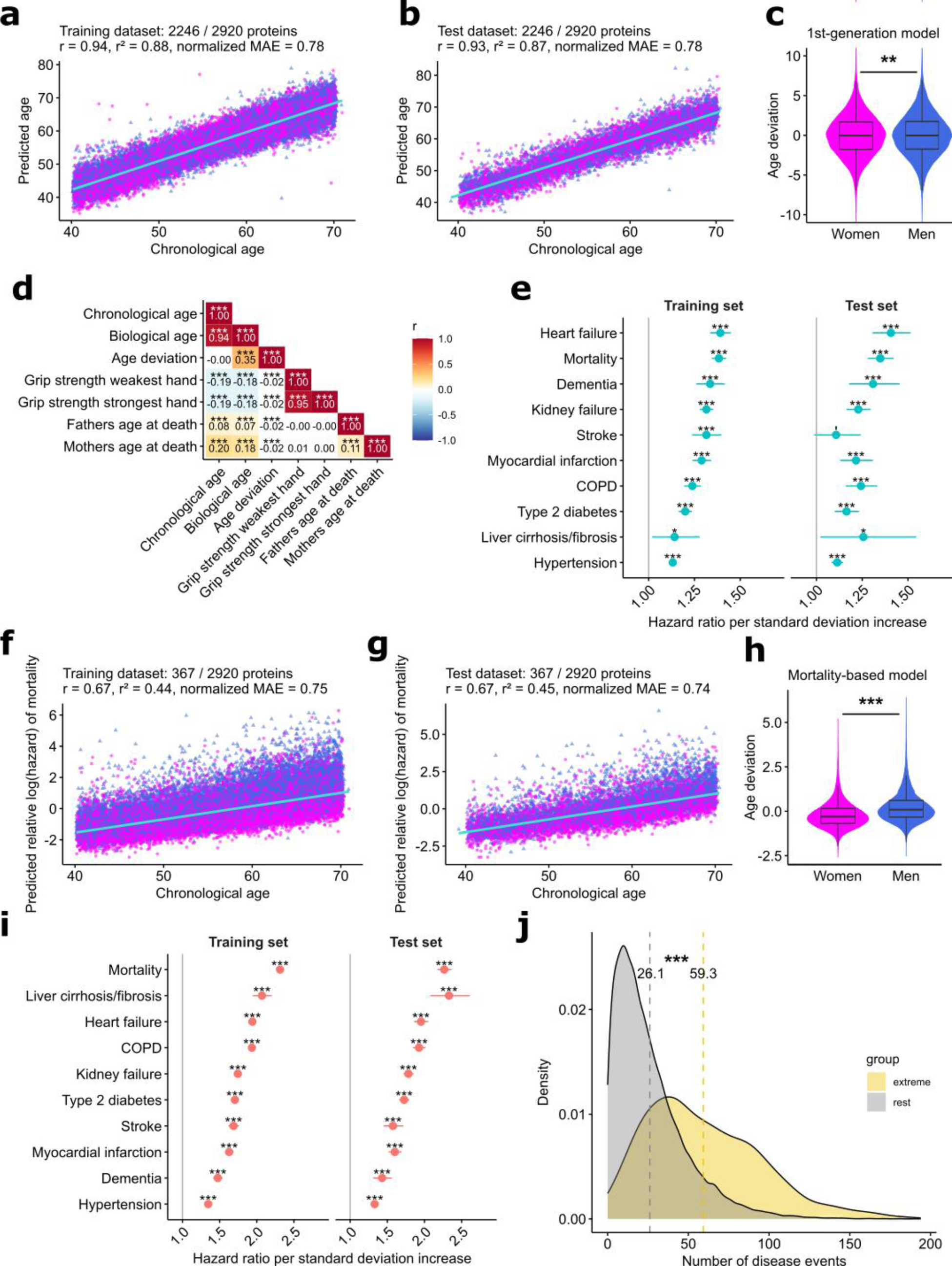
Plasma proteome-based models of biological age. **a – b.** Biological age as predicted by our 1^st^-generation proteome aging model has a strong positive correlation with chronological age in training (*n* = 42,412 UK Biobank participants) (a) and test datasets (*n* = 10,603 UK Biobank participants) (b). The number of proteins with non-zero coefficients is shown as a fraction of the total number of proteins on which the models are trained. r: correlation coefficient, r²: coefficient of determination, normalized MAE: mean absolute error of the normalized residuals, magenta: women, blue: men. Robust regression lines with 95% confidence bands (shaded area) are added. **c.** Violin plot showing that age deviation is significantly lower in women (*n* = 28 580) vs men (*n* = 24 435). ‘**’: p = 0.007, Welch t-test. **d.** Heatmap showing the correlation coefficients (r) between biological age as predicted by our aging model, age deviation, and some continuous traits in the test dataset. Note that our model captures additional information besides chronological age, because age deviation correlates negatively with both grip strength and parental age of death. **e.** Estimated hazard ratios per unit of standard deviation in the chronological aging model are significantly larger than 1 for various common age-related diseases and all-cause mortality in training and test datasets, after correcting for chronological age, sex, and their interaction. **f – g.** Predicted relative log(hazard) of mortality correlates positively with chronological age in training (*n* = 42,412 UK Biobank participants, panel a) and test datasets (*n* = 10,603 UK Biobank participants, panel b). The number of proteins with non-zero coefficients is shown as a fraction of the total number of proteins on which the models are trained. r: correlation coefficient, r²: coefficient of determination, normalized MAE: mean absolute error of the normalized residuals, magenta: women, blue: men. Robust regression lines with 95% confidence bands (shaded area) are added. **h.** Violin plot showing that the age deviation of the mortality-based aging model is significantly lower in women (*n* = 28,580) vs men (*n* = 24,435). ‘***’: p < 1*10^−16^, Welch t-test. **i.** Estimated hazard ratios per unit of standard deviation in the mortality-based aging model are significantly larger than 1 for various common age-related diseases and all-cause mortality in training and test datasets, after correcting for chronological age, sex, and their interaction. ‘***’: BH-FDR < 0.001. **j.** Based on the mortality-based aging model, extreme agers (*n* = 359) have significantly more first occurrences of International Classification of Diseases, 10^th^ edition (ICD-10) disease annotations as compared to other UK Biobank participants (*n* = 52,938). p < 1*10^−16^, Welch t-test. Dotted lines show the mean number of ICD-10 disease codes annotated for both groups in UK Biobank category 1712. **d, e, i**: ‘’’: Benjamini-Hochberg false discovery rate (BH-FDR) < 0.1; ‘*’: BH-FDR < 0.05; ‘**’: BH-FDR < 0.01; ‘***’: BH-FDR < 0.001.

Our 1^st^-generation chronological aging model reveals a subtle but significant sex difference, with men predicted to be older than women (Fig. 1c). Predicted age correlates negatively with grip strength in both dominant and non-dominant hands (Fig. 1d), reproducing a well-known age-related decline. Age deviation, the residual from regressing predicted biological age on chronological age, also correlates negatively with grip strength, demonstrating the ability of our model to capture effects beyond chronological age. Parental age of death correlates positively with both chronological and protein predicted age (Fig. 1d). This is expected because the broad-sense heritability of human longevity is ∼15 – 30% (Ruby et al., 2018). Therefore, older individuals are more likely to have longer-living parents due to survivorship bias. However, age deviation correlates negatively with parental age of death, indicating that biologically younger individuals have on average longer-living parents. This finding shows that our aging model also captures heritable determinants of longevity. Although trained on chronological age, increased age deviation as predicted by our model significantly increases the hazard for a plethora of age-related diseases, such as heart failure, dementia, and stroke, as well as for all-cause mortality, even after correcting for chronological age and sex (Fig. 1e).

### Mortality-based models of aging provide insights into age-related diseases and capture differences among sexes

We also trained an aging model on the same dataset using Cox proportional hazard elastic net regression with mortality as the outcome (see Methods, Supp. Table 2). Despite chronological age not being included as the training outcome, our mortality-based model retains a reasonably strong correlation with chronological age, both in training (Fig. 1f, r = 0.67) and test datasets (Fig. 1g, r = 0.67). We validated both the 1^st^-generation aging model and the mortality-based model in the external Multi-Ethnic Study of Atherosclerosis (MESA) study described in Bild et al. (2002), in which the plasma proteomes of 921 participants of diverse ancestries (including White, African-American, Hispanic, and Asian) were collected at their first and fifth visits and quantified with the SomaScan platform (Gold et al., 2010). Despite the fact that our models were trained on Olink data and that only 27% of proteins from the 1^st^-generation aging model and 34% of the proteins from the mortality-based aging model were identified, both aging models retain strong correlation with aging (r = 0.57 and 0.56 for the 1^st^-generation aging model at the first and fifth visits, respectively, and r = 0.41 and 0.46 for the mortality-based aging model at the first and fifth visits, respectively, Supp. Fig. 1).

Our mortality-based model finds a more pronounced difference in biological age between men and women than the chronological aging model, reflecting the overall higher mortality risk for men vs women (Crimmins et al., 2019) (Fig. 1h). Trained with mortality as an outcome, this model not only performs better at predicting the hazard for all-cause mortality, but also for various age-related diseases (Fig. 1i). Note that with hazard ratios of mortality of 1.38 and 1.35 per unit increase of standard deviation in training and test sets, respectively, our chronological aging model performs on par with established biomarkers of aging such as PhenoAge (Levine et al., 2018) and extrinsic epigenetic age acceleration (EEAA, based on (Hannum et al., 2013)) (Hillary et al., 2020). With hazard ratios of mortality equaling 2.31 and 2.27 per unit increase in standard deviation in training and test sets, our mortality-based model outperforms even the strongest biomarkers of mortality, including GrimAge (Lu et al., 2019), with its hazard ratio ∼1.3 – 1.9 per unit increase in standard deviation (Moqri et al., 2024).

We further assessed how the mortality-based model could be informative at the level of an individual. We identified participants with significantly increased age deviation (“extreme agers”, see Methods). Interestingly, these participants have significantly more disease diagnoses compared to the rest of the population (Fig. 1j).

### Plasma proteomics reveals markers of aging and mortality

We assessed the associations of each of the individual 2,923 measured proteins with chronological age, adjusting for sex. We found 1,664 proteins with significant positive associations with chronological age, and 665 proteins with significant negative associations (Fig. 2a, Supp. Table 3). Some interesting hits include GDF15 and NEFL. GDF15 is a mitokine that protects against aging-mediated inflammation (Moon et al., 2020) and was shown to be a robust predictor of all-cause mortality and various diseases in the UK Biobank (You et al., 2023). Neurofilament light chain (NEFL) is also known to increase with age and a known marker of mortality (Kaeser et al., 2021). These proteins were also identified as signatures of dementia (Guo et al., 2024). Gene set enrichment analysis reveals significant increases in proteins involved in biological adhesion and locomotion with chronological age (Fig. 2b). Interestingly, these proteins also strongly contribute to both our chronological age-based and mortality-based models.

**Figure 2.**
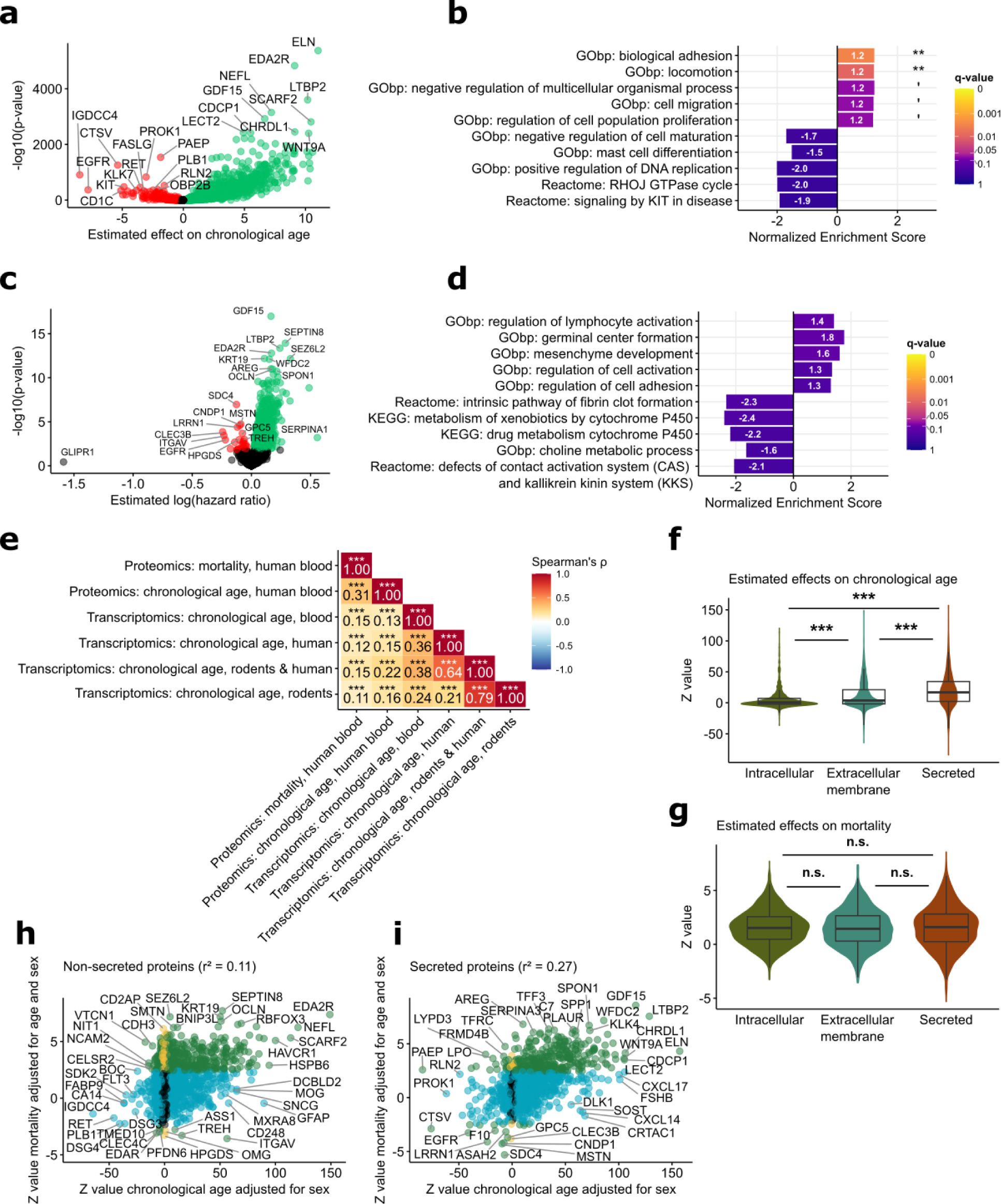
Proteomic signatures/biomarkers of chronological age and mortality. **a.** Volcano plot showing the effects on chronological age after correction for sex for 2,923 proteins in 53,016 UK Biobank participants. Increased abundance of 1,664 proteins (green dots) is significantly linked to higher chronological age, while increased abundance of 665 proteins (red dots) is significantly linked to lower chronological age at a 5% Benjamini-Hochberg false discovery rate (BH-FDR) significance threshold. **b.** Five most significantly up- and downregulated gene sets after gene set enrichment analysis on the effect on chronological age. Proteins in the gene sets “biological adhesion” and “locomotion” are significantly increased with increased chronological age (BH-FDR-adjusted q-values 0.003 and 0.009, respectively). **c.** Volcano plot showing the log(hazard ratio) of dying after correction for chronological age, sex and their interaction for 2,923 proteins in 53,016 UK Biobank participants. Increased abundance of 892 proteins (green dots) is significantly linked to increased risk of dying, while increased abundance of 27 proteins (red dots) is significantly linked to decreased risk of dying at a 5% Benjamini-Hochberg false discovery rate (BH-FDR) significance threshold. **d.** Five most significantly up- and downregulated gene sets after gene set enrichment analysis on the log(hazard ratio) of dying. No gene sets are significant at a 5% BH-FDR significance threshold. **e.** Heatmap showing the correlation coefficients (r) between normalized enrichment scores of pathways associated with blood proteomic signatures of chronological age and mortality, and blood and multi-tissue transcriptomic signatures of chronological age derived from Tyshkovskiy et al. (2023). **f.** Violin plots showing the Z values for the effects on chronological age after correction for sex for intracellular proteins, proteins located at the extracellular membrane, and secreted proteins. Plasma concentrations of secreted proteins are significantly increased with age as compared to intracellular and plasma membrane proteins, while extracellular membrane proteins are also significantly increased with age as compared to intracellular proteins (single-step adjusted p-values < 1.5*10^−10^). **g.** Violin plots showing the Z values for the hazard of dying after correction for chronological age, sex and their interaction, for intracellular proteins, proteins located at the extracellular membrane, and secreted proteins. The Z values are not significantly different between the three groups of proteins. **h – i.** Correlations between Z values for the effects on chronological age after correction for sex and Z values for the hazard ratio of dying after correction for chronological age, sex and their interaction, for non-secreted (g) and secreted (h) proteins. Cyan: proteins with significant effects on chronological age, green: proteins with significant effects on the hazard of dying, yellow: proteins with both significant effects on chronological age and significant effects on the hazard of dying. Significance thresholds were set at 5% BH-FDR. ‘’’: BH-FDR < 0.1; ‘*’: BH-FDR < 0.05; ‘**’: BH-FDR < 0.01; ‘***’: BH-FDR < 0.001.

We also assessed the associations of each of these individual plasma proteins with mortality using Cox proportional hazards models, while correcting for chronological age, sex, and their interaction. We found 892 proteins to be significantly associated with increased risk of all-cause mortality, while 27 proteins are significantly associated with decreased mortality risk (Fig. 2c, Supp. Table 4). Enrichment analysis did not reveal any gene sets to change significantly with mortality (Fig. 2d).

We then compared our proteomic signatures of aging and mortality with the corresponding transcriptomic aging signatures from Tyshkovskiy et al. (2023). Proteomic and transcriptomic signatures of chronological aging demonstrated statistically significant positive correlations (Supp. Fig. 2). The mortality proteomic signature also showed positive correlation with almost all transcriptomic signatures, albeit weaker. Remarkably, at the level of enriched pathways, proteomic and transcriptomic signatures demonstrated even stronger, significant correlations (Fig. 2e). Positive associations were mainly driven by upregulation of genes associated with inflammation, interleukin signaling, interferon signaling, the p53 pathway and apoptosis. This suggests that aging is associated with similar underlying biological processes across different molecular strata.

We further find that extracellular membrane proteins increase on average more significantly with age than intracellular proteins, and that secreted proteins increase on average more significantly than extracellular membrane proteins and intracellular proteins (Fig. 2f). Conversely, there is no statistically significant difference in the associations with mortality between these three groups of proteins (Fig. 2g). This indicates that the chronological aging signature is enriched for proteins that are secreted or expressed on the extracellular membrane, reflecting age-associated changes in these proteins, whereas the mortality signature elevated the proportion of intracellular proteins, likely reflecting increased cell death that releases these proteins into the plasma.

We next examined correlations between the associations of proteins with chronological age and mortality separately for secreted (Fig. 2h) and non-secreted proteins (Fig. 2i). We find that the correlation between aging and mortality prediction is much stronger for secreted than for non-secreted proteins (coefficient of determination r² = 0.27 for secreted proteins vs r² = 0.11 for non-secreted proteins). This is likely due to the presence of a significant amount of non-secreted proteins that do not change with age, but are still indicative of mortality when detected in aberrant amounts in the blood. Conversely, proteins that are secreted in different amounts with aging tend to also change in the same way.

### Organ-specific models capture diseases in these organs

We trained organ-specific models to predict chronological age (using elastic net) and mortality (using Cox elastic net) as outcomes (see Supp. Table 1, 2 for the model coefficients). For this, we followed the approach of Oh et al. (2023), and considered a protein to be organ-enriched if its Genotype-Tissue Expression (GTEx) expression (Lonsdale et al., 2013) is at least four times higher in one organ compared to all other organs (see Methods). We first assessed their association with chronological age (Fig. 3a, b). We chose to retain only organ-specific models that sufficiently captured features of aging, excluding models without a consistent coefficient of determination (r²) with chronological age above 10%.

**Figure 3.**
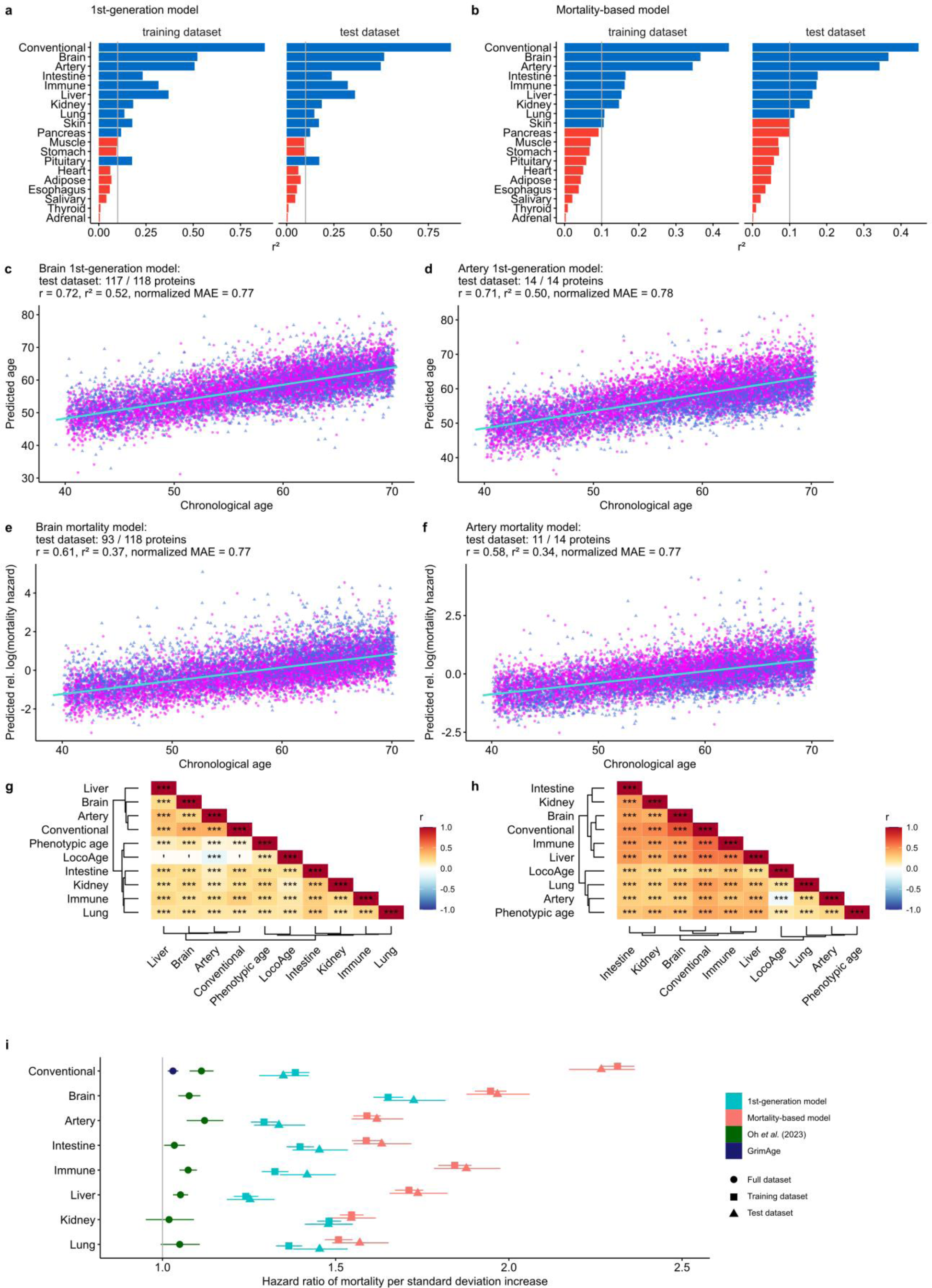
Organ-specific aging models predict chronological age and mortality. **a – b.** Barplots showing the coefficients of determination (r²) with chronological age for all organ-specific aging models and the conventional model in training (*n* = 42,412 UK Biobank participants) and test (*n* = 10,603 UK Biobank participants) datasets for the 1^st^-generation model trained on chronological age (a) and the mortality-based model (b). r² > 0.1 are colored in blue, r² < 0.1 in red. **c – d.** Biological age as predicted by our heart- (c), lung- (d) and immune-specific (e) 1^st^-generation aging models correlates positively with chronological age in the test dataset (*n* = 10,603 UK Biobank participants). The number of proteins with non-zero coefficients is shown as a fraction of the total number of proteins on which the models are trained. r: correlation coefficient, r²: coefficient of determination, normalized MAE: mean absolute error of the normalized residuals, magenta: women, blue: men. Robust regression lines with 95% confidence bands (shaded area) are added. **e – f.** Predicted relative log(hazard) of mortality correlates positively with chronological age in the test dataset (*n* = 10,603 UK Biobank participants) for the heart- (c), lung- (d) and immune-specific (e) mortality-based aging models. The number of proteins with non-zero coefficients is shown as a fraction of the total number of proteins on which the models are trained. r: correlation coefficient, r²: coefficient of determination, normalized MAE: mean absolute error of the normalized residuals, magenta: women, blue: men. Robust regression lines with 95% confidence bands (shaded area) are added. **g – h.** Heatmaps showing the correlation coefficients (r) for the age accelerations based on the 1^st^-generation (a) and mortality-based (b) organ-specific models and the conventional model (*n* = 53,015 UK Biobank participants), as well as the age deviations based on phenotypic age (*n* = 44,901 UK Biobank participants) and LocoAge (*n* = 10,428 UK Biobank participants). **i.** Hazard ratios per unit of standard deviation for the 1^st^-generation (cyan) and mortality-based (red) models, as well as for the Oh et al. (2023) study in the LonGenity cohort (green) and for DNAm-based GrimAge (Lu et al., 2019) (purple). ‘’’: BH-FDR < 0.1; ‘*’: BH-FDR < 0.05; ‘**’: BH-FDR < 0.01; ‘***’: BH-FDR < 0.001.

As organ-specific models are trained on only a subset of organ-specific proteins, their correlation with chronological age is generally weaker than the conventional model with, for example, coefficients of determination ranging from 0.58 to 0.72 in the test dataset for the brain and artery 1^st^-generation and mortality-based models (Fig. 3c – f). Plots for the other organs for both the training and test dataset can be found in Supp. Fig. 3-6. The conventional 1^st^-generation model, as well as the 1^st^-generation brain-, artery-, and kidney-specific models perform slightly better at predicting chronological age than the models of Oh et al. (2023), while other models perform slightly worse (Supp. Fig. 7). Biological ages, as predicted by these models, show significant positive correlations for all organs with each other and with chronological age (Supp. Fig. 8). Age deviation also correlates strongly for organ-specific models, reflecting coordinated aging across organs and systems (Fig. 3g,h). Age deviations for all these models also correlate positively with phenotypic age as defined by Levine et al. (2018), and with biological age calculated based on locomotor activity, as defined by Pyrkov et al. (2018), with the exception of the artery model. Thus, organ-specific models also align with physiological clocks, consistent with systemic organismal aging.

Overall, our 1^st^-generation organ-specific models perform in a range similar to the models from the Oh et al. (2023) study in predicting all-cause mortality, with our brain-, intestine-, kidney- and lung-specific models performing better, and the other organ-specific models performing worse (Fig. 3i). However, our mortality-based organ-specific models strongly outperform the 1^st^-generation organ-specific models from the Oh et al. (2023) study (Fig. 3i). All our models also strongly outperform GrimAge, a DNA methylation clock trained on 88 plasma proteins and smoking pack-years (Lu et al., 2019).

We examined if these organ-specific models could predict organ-specific diseases. Strikingly, organ-specific aging was indeed associated with increased risk of organ-specific diseases. For example, the hazard ratio for the mortality-based liver aging model is highest for liver cirrhosis/fibrosis, followed by kidney failure and heart failure, and these are higher than the hazard ratios for mortality on which the mortality-based heart aging model was trained (Fig. 4a). The kidney-specific model performs the best at reporting liver cirrhosis/fibrosis and kidney failure, followed by type 2 diabetes and heart failure, and only then mortality (Fig. 4b), while the lung-specific model best predicts chronic obstructive pulmonary disease (COPD) and only then mortality (Fig. 4c).

**Figure 4.**
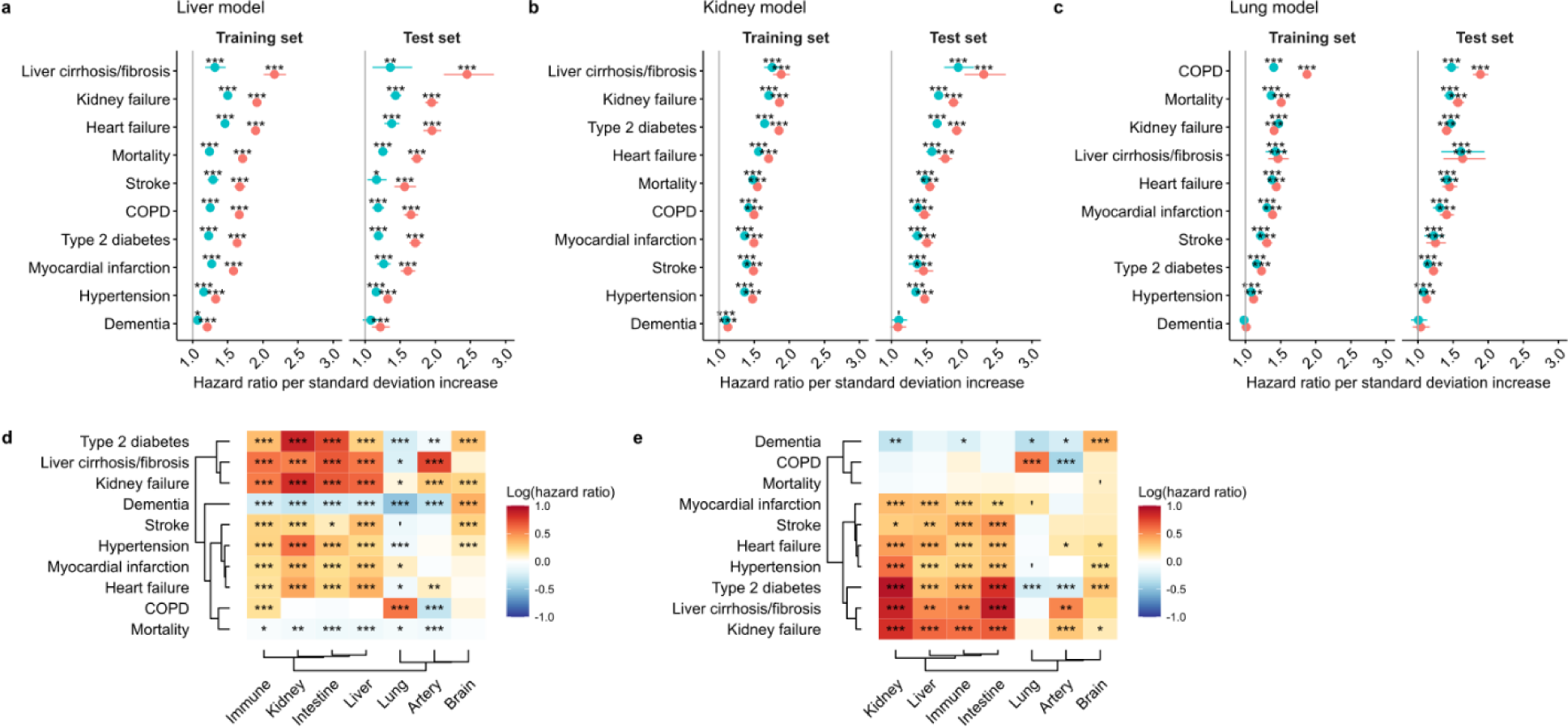
Organ-specific aging models predict organ-specific diseases. **a – c.** Hazard ratios per unit of standard deviation in the 1^st^-generation (cyan) and mortality-based (red) liver (a), kidney (b), and lung (c) aging models for various common age-related diseases and all-cause mortality in training (*n* = 42,412 UK Biobank participants) and test (*n* = 10,603 UK Biobank participants) datasets. **d – e.** Heatmaps showing log-hazard ratios per unit of predicted log(mortality hazard) in the mortality-based organ-specific models for various common age-related diseases and all-cause mortality in training (*n* = 42,412 UK Biobank participants) and test (*n* = 10,603 UK Biobank participants) datasets, after correcting for conventional mortality-based biological age, chronological age, sex, and the interaction between chronological age and sex. ‘’’: BH-FDR < 0.1; ‘*’: BH-FDR < 0.05; ‘**’: BH-FDR < 0.01; ‘***’: BH-FDR < 0.001.

When assessing the effects of the mortality-based organ-specific models on age-related diseases after correcting for conventional model’s mortality-based biological age, as well as for chronological age, sex, and their interaction, we uncovered strong relations between organ-specific aging and organ-specific diseases. For example, increased age as predicted by the mortality-based lung model is a predictor of chronic obstructive pulmonary disease (COPD), independent of biological age predicted by the conventional model (Fig. 4d, e). Similarly, the brain model is the only organ-specific model for which increased organ age is indicative of dementia (Fig. 4d, e), whereas the kidney model is the strongest predictor for type 2 diabetes and kidney failure. These findings demonstrate that organ-specific models capture additional features of organ-specific diseases that are not present in the conventional model, and more broadly they reflect diseases as accelerated aging of the organs (or systems) that harbor these diseases.

### Organ-specific aging is associated with environmental factors

We assessed how one’s field of work associates with the biological age predicted by our models. Individuals with professions that typically require higher levels of education, such as scientists, managers, teachers, and medical doctors, have significantly lower age deviations as measured by the mortality-based aging model, and this is observed in most organs (Fig. 5a). Conversely, professions requiring repetitive work such as cleaning workers, factory workers, and telephone operators have significantly higher age deviations. In line with this finding, it has been shown that high levels of autonomy and high social status at the workplace are associated with greater health, while repetitive low-paid jobs are associated with poorer health (Burgard & Lin, 2013). These effects are also captured by our mortality-based aging models.

**Figure 5.**
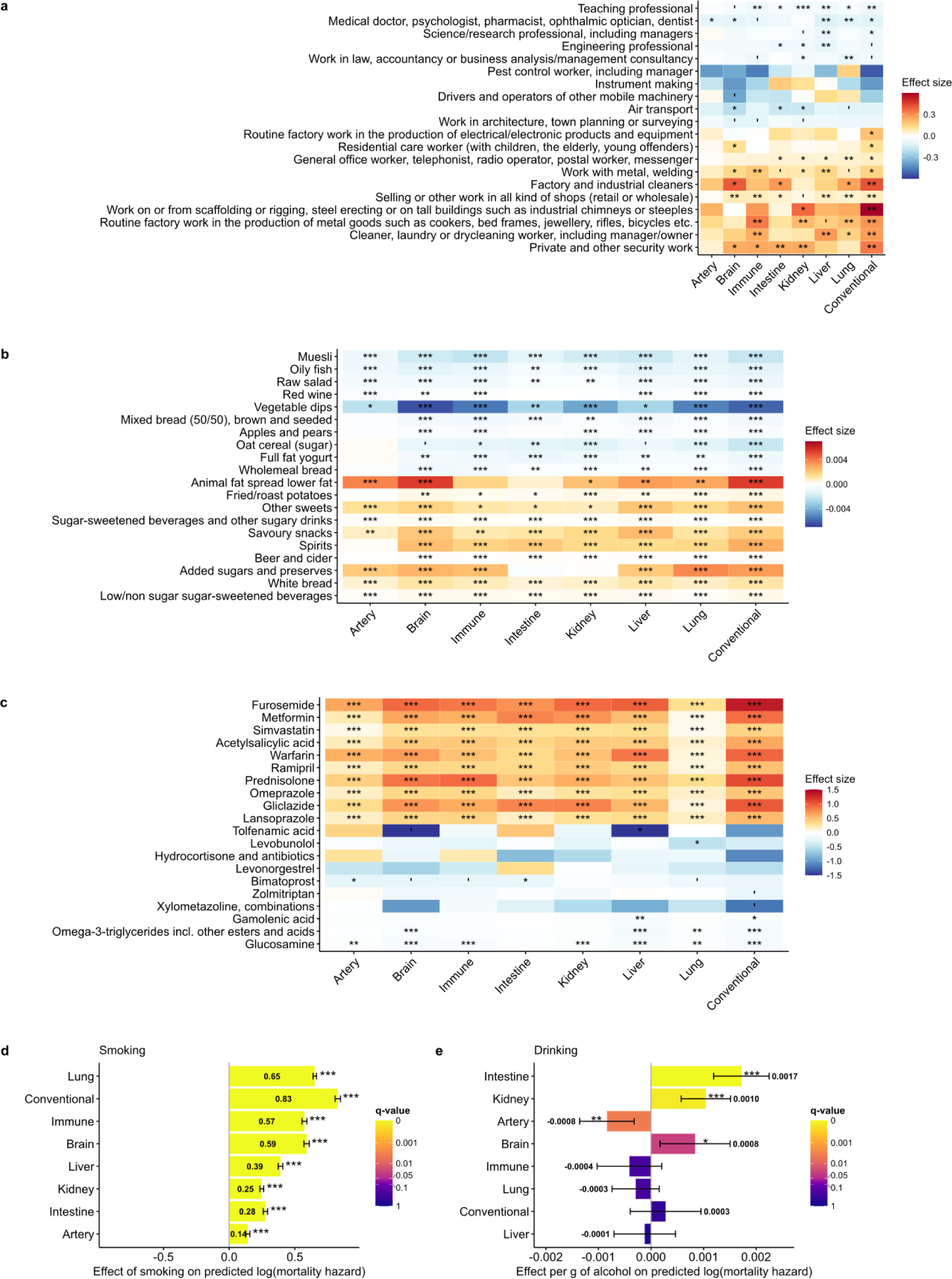
Organ-specific aging is associated with environmental factors. **a.** Heatmap showing the effects on the organ-specific mortality-based aging models for the professions with the most significant positive and negative associations (10 each) with age deviation, as measured by the conventional mortality-based aging model on *n* = 53,015 UK Biobank participants. **b.** Heatmap showing the effects on the organ-specific mortality-based aging models for the food groups with the most significant positive and negative associations (10 each) with age deviation, as measured by the conventional mortality-based aging model on *n* = 53,015 UK Biobank participants. **c.** Heatmap showing the effects on the organ-specific mortality-based aging models for the medications with the most significant positive and negative associations (10 each) with age deviation, as measured by the conventional mortality-based aging model on *n* = 53,015 UK Biobank participants. ‘’’: BH-FDR < 0.1; ‘*’: BH-FDR < 0.05; ‘**’: BH-FDR < 0.01; ‘***’: BH-FDR < 0.001. **d.** Barplots showing the differences in log(hazard of mortality) as predicted by the organ-specific mortality-based aging models between current smokers and participants who never smoked. Whiskers denote 95% confidence intervals (*n* = 53,015 UK Biobank participants). **e.** Barplots showing the effects of one additional g of alcohol intake on the log(hazard of mortality) as predicted by the organ-specific mortality-based aging models. Whiskers denote 95% confidence intervals (*n* = 53,015 UK Biobank participants). All analyses in this figure are based on ordinary least squares regressing biological age on the predictor of interest while correcting for chronological age, sex and their interaction. ‘’’: BH-FDR < 0.1; ‘*’: BH-FDR < 0.05; ‘**’: BH-FDR < 0.01; ‘***’: BH-FDR < 0.001.

We also assessed the associations of diet with biological age as measured by our mortality-based models (Fig. 5b). UK Biobank e-mailed online 24-hour recall dietary questionnaires to participants after their first visit. Using this information as a proxy for dietary habits, we assessed the associations of 93 different food groups on biological age as predicted by our organ-specific aging models, after correction for sex, biological age and their interaction. Foods which are generally considered healthy such as apples and pears, and raw salads, as well as foods associated with higher socioeconomic status, such as red wine consumption are associated with lower biological age. Unhealthy foods, such as added sugars, processed meat, spirits, beer and cider are associated with increased biological ages as measured by the mortality-based model.

We also assessed the associations of 582 medications with the biological ages predicted by our models after correction for sex, biological age and their interaction (Fig. 5c). Furosemide, a drug that is used to treat oedema in acute life-threatening conditions such as heart failure, liver cirrhosis, kidney failure and nephrotic syndrome, is most strongly associated with high biological age using the conventional model. Interestingly, the use of drugs such as simvastatin, used against high cholesterol and high triglyceride levels, and metformin, used to treat type 2 diabetes, are also strongly associated with increased biological age, likely reflecting diseased status of participants. Conversely, glucosamine, omega-3-triglycerides and gamolenic acid, which are also available as food supplements, are significantly associated with lower biological age. This might be related to higher socioeconomic status and generally healthier lifestyle of the people who use these supplements.

Finally, we investigated the impact of smoking and drinking alcohol on organ-specific aging. Even though the average effect size is the largest for the conventional model, with a -log10(p-value) of 1478, the difference in predicted log(hazard of mortality) between current and non-smokers is the most significant for the lung model, followed by the immune model (-log10(p-value) = 559) and the conventional model (-log10(p-value) = 536) (Fig. 5d). For drinking alcohol, we notice the largest increase in predicted log(mortality hazard) in the intestine model, followed by the kidney and brain models (Fig. 5e). Interestingly, drinking alcohol has a significant negative effect on the log(mortality hazard) predicted by the artery model (Fig. 5e).

### Longitudinal models of aging predict future health outcomes

We leveraged longitudinal data in UK Biobank to assess the effects of changes in diets and medications on the predicted biological age. UK Biobank has unique follow-up plasma proteomics data for 1,132 participants at their third visits, and for 1,006 participants at their fourth visits. For the 1,006 participants with proteomics data available at visit 1, 3, and 4, we fit the mortality-based aging model on the 1,461 available proteins (Supp. Fig. 9, Supp. Table 5). We used a mixed-effects model (see Methods) to estimate the rate of aging of these participants. As expected, our model shows that 976 out of 1,006 participants (97%) increase in biological age over time (Fig. 6a), and this is also the case for organ-specific longitudinal models (Supp. Fig. 10).

**Figure 6.**
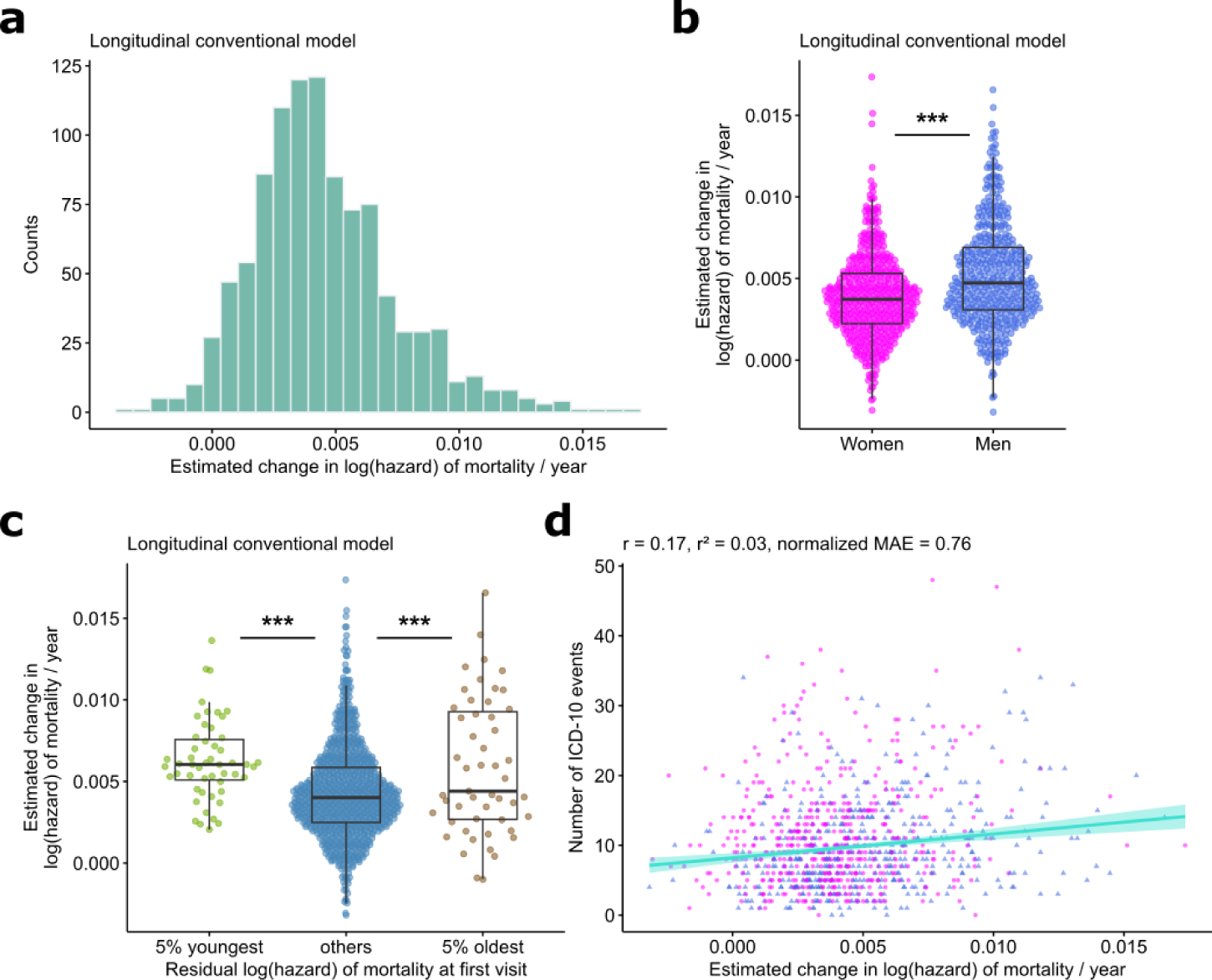
Longitudinal aging models. **a.** 976 out of *n* = 1,006 individuals increase in biological age. The histogram shows the distribution of the slopes of the biological ages as predicted by the conventional mortality-based model for the *n* = 1,006 individuals that have proteomics data available for their first, third and fourth visits. **b.** Men increase faster than women in biological age (p = 2*10^−12^, Welch t-test). **c.** The 5% youngest and 5% oldest age faster than the 90% in the middle (single-step-adjusted p = 1*10^−5^ and 8*10^−4^ respectively). **d.** An increase of 0.01 units in the change of log(hazard) of mortality per year as estimated by our conventional longitudinal aging model results in an increase of on average 4 extra ICD-10 disease annotations (ordinary linear regression, p = 1*10^−7^, *n* = 1,006 UK Biobank participants). r²: coefficient of determination, normalized MAE: mean absolute error of the normalized residuals, magenta: women, blue: men. Robust regression lines with 95% confidence bands (shaded area) are added.

Interestingly, not only are men on average biologically older than women (Fig. 1c, Fig. 1h), they also seem to age faster (p = 3*10^−11^, Fig. 6b). We investigated how the relative biological age at the first visit impacts the rate of aging as measured by our models. We find accelerated aging in both the cohorts with the lowest and the highest biological ages relative to their chronological age (Fig. 6c, Supp. Fig. 11). The accelerated aging of biologically young individuals can be explained by regression to the mean. However, the accelerated aging of the biologically oldest cohort suggests that our model effectively captures biological markers of increased mortality, and that these biologically older individuals are more likely to continue deteriorating in health. Indeed, an individual with a slope of 0.01 has on average 4 extra ICD-10 disease annotations as compared to an individual with slope 0 (Fig. 6d).

### Organ-specific models provide insights into organ-specific diseases in independent datasets

We assessed the performance of our aging models in an independent Filbin et al. (2021) dataset, which consists of 306 COVID-positive and 78 COVID-negative individuals for which the Olink plasma proteins were measured at days 0, 3, and 7 and the acuity levels were recorded days at 0, 3, 7, and 28.

The conventional mortality model strongly predicts chronological age (r = 0.62, Supp. Fig. 12a), but the lung mortality model has a slightly negative association with chronological age (Supp. Fig. 12b). Conversely, while the conventional model does not differentiate COVID positive from COVID negative individuals (Fig. 7a), the lung model makes a strong significant distinction between both groups (Fig. 7b, difference in average age deviation: 0.05, p = 7*10^−14^). However, while the age deviation predicted by the conventional model is predictive of disease severity (p = 3*10^−^13, r = 0.36, Fig. 7c), the lung model shows no significant differences according to severity of COVID infection (p = 0.5, r = 0.03, Fig. 7d). These observations are in line with the fact that the lung is an obvious entry organ for the SARS-CoV2 virus, turning the lung model into an accurate predictor for COVID status. However, severe COVID is a systemic disease, and hence the severity of the disease can be better assessed with a systemic conventional model. Indeed, all organ-specific models except for the lung-specific model are also predictive of COVID severity (Supp. Fig. 13).

**Figure 7.**
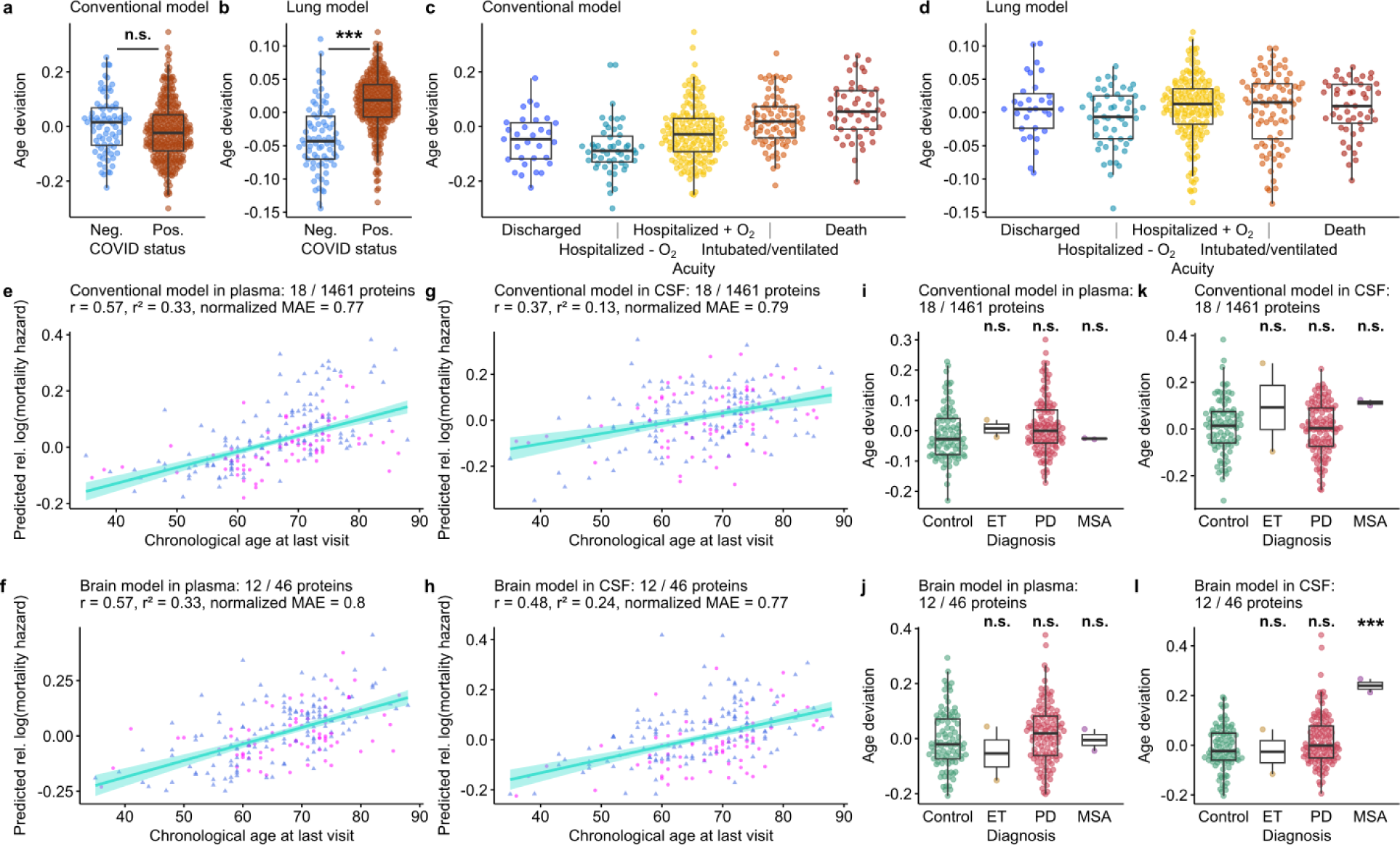
Organ-specific aging models reveal the relationship between aging and disease. **a.** Age deviation as predicted by the conventional model does not significantly differ between COVID negative (*n* = 78) and positive (*n* = 305) individuals at day 0 (p = 0.07, Welch t-test). **b.** Age deviation as predicted by the lung model is significantly higher in COVID-positive individuals (*n* = 305) compared to COVID-negative individuals (*n* = 78) (p = 7*10^−14^, Welch t-test). **c.** Increased age deviation as predicted by the conventional model is significantly associated with increased COVID infection severity (ordinary least squares p-value = 3*10^−13^, r = 0.36). **d.** Age deviation as predicted by the lung model is not significantly associated with COVID infection severity (ordinary least squares p-value = 0.5, r = 0.03). **e – f.** Predicted relative log(hazard) of mortality as predicted by the conventional (e) and brain (f) aging models correlates positively with chronological age in plasma samples (*n* = 212 individuals). The number of proteins with non-zero coefficients is shown as a fraction of the total number of proteins on which the models are trained. r: correlation coefficient, r²: coefficient of determination, normalized MAE: mean absolute error of the normalized residuals, magenta: women, blue: men. Robust regression lines with 95% confidence bands (shaded area) are added. **g – h.** Same as for (e – f), but applied to CSF (*n* = 212 individuals). **i.** In plasma, the conventional aging model does not detect significant differences in age deviation in the essential tremor (ET) group (*n* = 2 individuals), the Parkinson’s disease (PD) group (*n* = 118 individuals), and the multiple systems atrophy (MSA) group (*n* = 2 individuals), as compared to the control group (*n* = 90 individuals). The number of proteins with non-zero coefficients is shown as a fraction of the total number of proteins on which the models are trained (single-step-adjusted p = 0.3, 1, and 1, respectively). **j.** Same as in (i), for the brain model. Single-step-adjusted p = 0.8, 0.8, and 1, respectively. **k.** In CSF, the conventional aging model does not detect significant differences in age deviation in the essential tremor (ET) group (*n* = 2 individuals), the Parkinson’s disease (PD) group (*n* = 118 individuals), and the multiple systems atrophy (MSA) group (*n* = 2 individuals), as compared to the control group (*n* = 90 individuals) (single-step-adjusted p = 0.7, 0.8, and 0.4, respectively). **l.** Same as in (k), for the brain model. However, for the brain model in CSF, the biological age of the MSA group is significantly increased compared to the control group (Single-step-adjusted p = 5*10^−5^). The differences in biological age for the comparisons ET vs control and PD vs control are not significantly different (Single-step-adjusted p 0.9, and 0.2, respectively). Data in (a – d) are from the Filbin et al. (2021) dataset, data in (e – l) are from the Dammer et al. (2022) dataset.

We further focused on plasma and cerebrospinal fluid (CSF) samples, wherein 1,536 proteins were measured with the Olink platform (Dammer et al. (2022)). Out of 212 participants, 90 were classified as controls, 2 as having essential tremor (ET), 118 as having Parkinson’s disease (PD), and 2 as having multiple system atrophy (MSA), a rare fatal progressive neurodegenerative disorder. Our conventional mortality-based aging model and brain-specific model both predict chronological age and perform on par in plasma (Fig. 7e, f, coefficients of determination r = 0.57 for both). However, in CSF, the brain model outperforms the conventional model (Fig. 7g, h, coefficients of determination r = 0.37 and 0.48 for the conventional and brain models, respectively).

We then compared our model trained on the UK Biobank plasma to 1^st^-generation conventional (Supp. Fig. 14) and brain-specific (Supp. Fig. 15) models trained directly on the CSF data from Dammer et al. (2022) (Supp. Table 6). As expected, the 1^st^-generation conventional model and brain-specific model perform better at predicting chronological age (Supp. Fig. 14a – c, Supp. Fig. 15a – c). The brain-specific CSF model, but not the conventional model, is able to differentiate PD and MSA from controls (single-step adjusted p = 0.01, and 0.002, respectively, Supp. Fig. 14d, Supp. Fig. 15d). In plasma, neither the conventional nor the brain-specific mortality-based model find significant differences in biological age between controls, essential tremor, Parkinson’s disease and MSA (Fig. 7i, j). Given its low accuracy in the CSF (r = 0.37), the conventional mortality-based model is again unable to detect a significant difference between control, ET, PD and MSA (Fig. 7k). However, our brain-specific mortality-based model trained on UK Biobank plasma is better at discerning MSA (Fig. 7l, single-step-adjusted p = 5*10^−5^). However, these results will need to be verified in samples with larger numbers of MSA cases. Finally, we used the data from Damsky et al. (2022) to assess our models in the context of sarcoidosis, a disease characterized by the growth of inflammatory granulomas throughout the body. We find that there is a significant increase in mortality hazard as predicted by the immune and intestine models (Hommel-adjusted p-values = 0.001 for both), but not the conventional model nor the other organ-specific models (Supp. Fig. 16).

## Discussion

The question of whether aging is a disease has long been debated. The discourse on the relationship between aging and disease has been centered on whether aging is a normal, natural, physiological process, or whether it represents a pathology (Gladyshev & Gladyshev, 2016). In our study, we turn the table by posing the question of whether chronic disease is a manifestation of aging. Our data on organ-specific aging patterns and their connections with diseases suggest that chronic age-related diseases represent the accelerated aging of respective organismal systems or subsystems.

It has been previously suggested that accelerated age-associated alterations within the heart and arteries should be regarded a type of cardiovascular disease (Lakatta, 2015). It was argued that the concept of cardiovascular aging as a disease should be incorporated into clinical medicine. Similarly, many of the changes observed in the lungs during aging, such as declined lung function, increased gas trapping, loss of lung elastic recoil, and enlargement of the distal air spaces, are also characteristic of COPD. These observations prompted researchers to consider COPD as a condition of accelerated lung aging (MacNee, 2016). Not surprisingly, type 2 diabetes has also been considered as a condition of accelerated aging (Kuss et al., 2024). However, viewing chronic disease as an accelerated aging condition does not necessarily mean that faster aging of an organ always results in a disease, nor does it imply that accelerated organ aging may be a pathogenic mechanism in all individuals with chronic disease. Indeed, not all very old organisms develop diseases in all organs. Disease might be better understood as an extreme form of aging within a system, which can occur at any age, but is more likely to occur later in life. In some instances, such extreme aging may affect only a subsystem within an organ, compromising its function, or it may span functions of interacting organs.

It is important to note that accelerated organ aging may be compensated for by interactions with younger organs and systems within the organism, although such compensation may decrease with aging. Moreover, “gravitation” of a system characterized by accelerated aging towards other systems within the organism that exhibit lower biological age would lead to a state of the accelerated aging system that does not parallel the one achieved by normal aging.

In our study, we leveraged large-scale plasma proteomics data from over 53,000 UK Biobank participants to develop chronological (1^st^-generation), mortality, longitudinal and organ-specific aging models. Our findings indicate that chronic diseases typically manifest as accelerated organ- and system-specific aging phenomena.

The rich proteomics data available in UK Biobank, which comprises measurements of almost 3,000 proteins in the plasma of over 50,000 individuals (Eldjarn et al., 2023), allows for the development of highly accurate aging biomarkers (Argentieri et al., 2023). Recent studies have demonstrated that epigenetic clocks trained on different tissues may yield varied outcomes, suggesting that organs may age at different rates (Prattichizzo et al., 2024). Building on the findings of Oh et al. (2023), who demonstrated that the secreted proteomes of different organs could age differently between individuals, organ-specific aging models have garnered increasing attention in aging research. Our study advances this concept, by utilizing the extensive UK Biobank dataset to develop robust organ-specific aging models with superior statistical power and generalizability.

We initially developed conventional aging models, trained on both chronological age (first-generation aging model) or mortality. Both models, especially the mortality-based aging model, demonstrated substantial predictive capability for future all-cause mortality, with the mortality-based model outperforming most, if not all, existing, validated biomarkers of aging (Moqri et al., 2024). Moreover, the hazard ratios of these models were also consistently high for a broad spectrum of severe age-related diseases.

We also demonstrated sex differences in the rate of biological aging, as evidenced by both the chronological-age-based model and the mortality-based model, which consistently showed that men have higher biological ages on average compared to women. Moreover, our longitudinal model also revealed that men tend to age at a faster rate than women. This finding aligns with other biomarkers of aging, which similarly indicate lower biological ages of women (Hägg & Jylhävä, 2021).

We proceeded to develop organ-specific models of aging, building upon the work of Oh et al. (2023). These researchers showed a correlation between accelerated organ aging and increased risk of mortality, as well as a link between organ-specific diseases and faster aging of those organs. Our findings indicate that organs and systems exhibit distinct characteristic aging signatures in the plasma proteome, which can be effectively captured by organ-specific aging models. Notably, despite being trained solely on chronological age or mortality data, our organ-specific models were predictive of organ-specific diseases.

For instance, the mortality-based liver aging model showed the highest hazard ratios for liver cirrhosis/fibrosis, followed by kidney failure and heart failure. Interestingly, kidney failure and heart failure often arise in liver cirrhosis patients (Xanthopoulos et al., 2019), with up to 50% of hospitalized patients with cirrhosis developing acute renal failure (Nadim & Garcia-Tsao, 2023). The lung aging model was the best predictor for chronic obstructive pulmonary disease (COPD), and, after correcting for conventional mortality-based age, the kidney-based model associated most strongly with renal failure and type 2 diabetes. The association between kidney aging and type 2 diabetes makes sense as the kidney is the most important target of microvascular damage in diabetes, and about half of all type 2 diabetes patients develop chronic kidney disease (Nadim & Garcia-Tsao, 2023).

Interestingly, while dementia is associated with accelerated brain aging, it corresponds to decelerated aging in other organs. This observation may be explained by the fact that dementia typically develops later than most other chronic diseases. It becomes prevalent primarily in extremely old individuals, who are expected to age slower to reach the ages when this disease develops. In contrast, individuals with accelerated aging of all organ systems or those with uniform organ aging would likely succumb primarily to diseases that manifest earlier in life. Taken together, these results suggest that age-related chronic diseases may arise from accelerated aging processes within specific organs or systems, rather than from a uniform, systemic aging process, and that our organ-specific aging models are able to predict the risk of age-related organ-specific diseases that is not fully captured by conventional aging biomarkers.

Our analyses demonstrated that many lifestyle factors, such as diet, occupation, and medication use, were strongly associated with biological age predicted by our models. Notably, individuals in professions requiring higher education and high autonomy tended to exhibit lower biological ages, while those in routine, low-paid jobs showed accelerated aging across multiple organs. Similarly, healthy dietary patterns, such as the consumption of fruits and vegetables, were associated with lower biological ages, whereas unhealthy foods, like processed meats and added sugars, were linked to increased biological aging.

We also observed some organ-specific associations of behavior with biological age. For example, since smoke directly enters the body through the lungs, it is expected that smoking primarily affects lung health. Indeed, the lung-specific model outperforms all other organ-specific models and the conventional model in distinguishing current smokers from non-smokers. Similarly, as alcohol is absorbed into the body through the intestine, the intestine-specific model shows the highest increase in biological age for each additional unit of alcohol consumed. While one needs to be wary of the many confounders associated with alcohol consumption in observational studies, it is nevertheless interesting to speculate that the vasodilating and damage buildup clearing properties of alcohol might contribute to the lower biological ages in the artery-specific model. There is indeed still no consensus regarding the effects of moderate alcohol consumption on cardiovascular disease specifically (Hoek et al., 2022).

We applied our organ-specific models in two independent datasets. In the COVID-19 dataset from Filbin et al. (2021), the lung-specific model accurately differentiated between COVID-positive and COVID-negative individuals, reflecting the lung as the primary entry point for SARS-CoV-2. Conversely, the conventional model better in predicting disease severity, underscoring the systemic nature of severe COVID-19 infection. Similarly, both the brain-specific and kidney-specific models could predict disease severity. Acute kidney injury is a common occurrence in COVID-19 patients (La Porta et al., 2022), and severe COVID has been linked to accelerated brain aging (Mavrikaki et al., 2022).

In the neurodegenerative disease dataset from Dammer et al. (2022), we revealed that the brain-specific models outperformed conventional models in predicting chronological age when applied to CSF data. Distinguishing PD patients and controls proved challenging. Only a brain-specific 1^st^-generation model trained on the CSF data itself was able to achieve significant differentiation between PD and controls (single-step-adjusted p-value = 0.01), but our mortality-based conventional and brain-specific models trained on UK Biobank plasma data failed to distinguish PD from controls. Conversely, our UK Biobank plasma-trained brain model outperformed the CSF-trained 1^st^-generation models in predicting multiple system atrophy (MSA). This difference in predictive power might be attributed to the fact that our models are trained on mortality as an outcome. Parkinson’s disease is associated with a near-normal life expectancy, while MSA is a progressive, fatal neurodegenerative disorder with a median survival of only 8.6 years after diagnosis (Figueroa et al., 2014). Although these results need to be confirmed in a dataset with a larger number of MSA cases, this observation seems to underscore the notion that aging models trained on mortality are more adept at predicting diseases with higher mortality risks.

Our study has several limitations. It is important to bear in mind that the UK Biobank data is inherently observational, and various observed and unobserved confounders can strongly influence the associations we observed. Furthermore, only a small fraction of the participants for whom longitudinal data is available were selected at random (*n* = 106). The other participants were enriched for numerous diseases that were of interest to UK Biobank’s Pharmaceutical partners. Hence, the longitudinal cohort is overall less healthy than the average UK Biobank participants, which can also introduce bias in the longitudinal analysis.

Overall, we have shown here the effectiveness of organ-specific aging models in capturing information beyond that captured by conventional aging models, and that this information is predictive of organ-specific disease. Based on these models, aging can impact different organs and systems at different rates. We further demonstrate that different lifestyles, including dietary habits, professional occupation and medication intake, are associated with varying rates of aging across different organs. These findings suggest that future anti-aging interventions should not only target organismal aging, but could also be tailored towards the oldest organs, calling for a more personalized approach towards anti-aging interventions. The concept of a chronic disease representing an accelerated aging of a specific system offers insights into the ongoing debate on whether aging itself should be considered a disease. Viewing the aging of a system or subsystem as reflective of a disease state suggests that aging of an entire organism could be viewed as a form of whole-organismal disease. However, this analogy is incomplete. Unlike accelerated aging at the system level that occurs non-autonomously in the background of interactions with younger systems, the aging of the whole organism occurs within a relatively isolated system.

## Methods

### Human cohorts

UK Biobank recruited ∼500,000 people aged between 40-69 years old from 2006 until 2010 from across the United Kingdom. This dataset contains detailed personal information and medical records, amongst others. Recently, (Eldjarn et al., 2023) used the high-throughput affinity-based proteomics Olink Explore 3072 platform to measure the levels of 3,072 proteins simultaneously in the plasma of 53,015 UK Biobank participants. They also measured the levels of 1,463 proteins in the plasma of 1,132 participants at the imaging visit (2014+) and for 1,006 participants at the first repeat imaging visit (2019+).

The datasets of Filbin et al. (2021) and Dammer et al. (2022) were used for validation purposes. Both studies measured 1,536 unique proteins with the Olink Explore 1536 platform. The Filbin et al. (2021) dataset contains 306 COVID-19 patients and 78 COVID-negative controls. The Dammer et al. (2022) dataset contains 118 PD patients, 2 ET patients, 2 MSA patients, and 90 controls.

### UK Biobank data processing

Grip strength of the strongest hand was defined as the highest grip strength between the left (UK Biobank field 46) and the right (UK Biobank field 47) hands, and grip strength of the weakest hand was defined as the lowest grip strength between both hands. Mother’s and father’s age of death were derived from fields 3526 and 1807, respectively. Participants with unknown ages of death for the parents, or those who replied “Yes”, “Prefer not to answer”, or “Do not know” to the question “Were you adopted as a child?” (field 1767) were excluded from the analyses related to parental age of death.

Mortality was recorded as the time of death, as stored in the death records (UK Biobank field 40000). Heart failure was defined as the first reported occurrence of heart failure (field 131354). Dementia was defined as the first reported occurrence of any of Alzheimer’s disease (field 130836), vascular dementia (field 130838), other dementia (field 130840), or unspecified dementia (130842). Kidney failure was defined as the first reported occurrence of any of acute renal failure (field 132030), chronic renal failure (field 132032), or unspecified renal failure (field 132034). Stroke was defined as the first reported occurrence of stroke, not specified as haemorrhage or infarction (field 131368). Myocardial infarction was defined as the first occurrence of acute (field 131298) or subsequent myocardial infarction (field 131300). Chronic obstructive pulmonary disease (COPD) was defined as the first reported occurrence of other chronic obstructive pulmonary disease (field 131492). Type 2 diabetes was defined as the first reported occurrence of non-insulin-dependent diabetes mellitus (field 130708). Liver cirrhosis/fibrosis was defined as the first reported occurrence of fibrosis and cirrhosis of liver (field 131666). Hypertension was defined as the first reported occurrence of any of essential (primary) hypertension (field 131286), or secondary hypertension (field 131294). Food intake was based on the total weight of food group yesterday (UK Biobank category 100118)(Perez-Cornago et al., 2021; Piernas et al., 2021). Job codings were obtained from job coding (field 22601) and, for each occupation, were assigned to that occupation if they have ever worked in it for at least 6 months for at least 15 hours per week. Smoking status was obtained from UK Biobank field 20116, collected at first visit.

Treatment/medication codes in the UK Biobank (field 20003) were streamlined and categorized through the following process. First, we applied mapping data generated by Wu et al. (2019) to map UK biobank medications taken by at least 10 subjects to their respective Anatomical Therapeutic Chemical (ATC) codes. ATC provides a classification system that enabled us to hierarchically categorize each UK medication into 5 distinct levels of classification (Anatomical Therapeutic Chemical Classification - Summary | NCBO BioPortal). We downloaded the ATC categories from Bioportal Bioontology, formatted them so each row corresponded to each of 5 classifications for a given drug, and then merged each drug with the UK biobank codes, creating a final table of medications with integrated UK biobank drugs and ATC classifications.

### Aging models

The Olink proteomics data was obtained from UK Biobank Record Table 1072. Proteins with > 50% missing values were removed from the analysis, retaining 2,920 out of 2,923 proteins. The proteomics data was then imputed using the k-nearest neighbors algorithm, with the default value of k = 10, as implemented in the impute.knn function from the impute R package v 1.68.0. The UK Biobank data (53,015 individuals) was then randomly split into a training (*n* = 42,412 individuals, 80% of the total) and a test dataset (*n* = 10,603 individuals, 20% of the total). In both training and test datasets, protein values were mean-centered, and the data was imported into Python.

Chronological aging (1^st^-generation) models were trained using elastic net regression on the plasma proteomics data, with chronological age as the outcome. Mortality-based models were trained using Cox proportional hazards elastic net regression with the time difference between death and the first visit as an outcome for the deceased subjects, and the time difference between the latest recorded death and the first visit as the outcome for the surviving subjects. Both models were optimized for the mean absolute error (MAE). These models were also used for the validation analysis on the SomaScan data in the Multi-Ethnic Study of Atherosclerosis (MESA) (Bild et al., 2002) dataset.

Normalized mean absolute errors (MAE) shown in the figures are the mean absolute errors calculated on the normalized residuals. Robust regression lines with 95% confidence bands shown in figures are based on robust linear regression (M estimation with Huber weights), as implemented by default in the rlm function from the MASS R package.

Extreme agers as shown in Fig. 1j were defined as follows. We normalized the age deviation (i.e. the residuals from an ordinary linear regression of predicted biological age on chronological age) by dividing them by the standard deviation of all age deviations. We then converted these normalized age deviations to p-values, assuming them to follow a standard normal distribution under the null hypothesis of no age deviation. We then converted the p-values to q-values using the Benjamini-Hochberg false discovery rate (FDR) method. Any participant with a q-value < 0.05 was considered to have a significant age deviation and therefore an extreme ager. We identified 359 of such individuals. First occurrences of ICD-10 disease codes were based on UK Biobank Category 1712, which contains information from primary care data, hospital inpatient data, death register records and self-reported medical condition codes mapped to 3-character ICD-10 codes.

### Annotation of proteins

To identify organ-enriched proteins, we incorporated information related to tissue transcription from the Genotype-Tissue Expression (GTEx) project (Lonsdale et al., 2013), in an approach similar to what has been done in Oh et al. (2023). We grouped the GTEx tissues into organs according to Supplementary Table 2 from these authors. A protein was then designated as organ-enriched if the average GTEx gene counts for an organ were at least four times higher than all other organs. We decided to make use of transcription data and not proteomics because transcription can inform about the organ of origin of a protein.

Organ-specific aging models were trained only on the subset of proteins enriched in a certain organ, in the same way as explained above for the conventional models. Finally, we only retained those organ-specific aging models for which the correlation coefficient (r) was consistently larger than 10% for both the chronological aging model and the mortality-based aging model, in both training and test sets.

For the analysis in Fig. 2f – g, we downloaded the human proteome from Uniprot on February 27, 2024. Proteins were considered secreted if they were annotated as belonging to the extracellular space in Gene Ontology cellular component (GOcc), or annotated as secreted in Uniprot’s subcellular location. Proteins were considered to belong to the extracellular membrane if they were not secreted and annotated as belonging to the plasma membrane in Gene Ontology cellular component (GOcc), or annotated as cell membrane in Uniprot’s subcellular location. All other proteins were considered to be intracellular. In Fig. 2h – i, secreted proteins are the same as for the analysis in Fig. 2f – g, and non-secreted proteins are all other proteins.

### Longitudinal analyses and external validation

For the longitudinal analyses, we only considered the 1,463 proteins that were measured in the Olink panels from the samples taken at the third and fourth visits. After filtering out proteins with >50% missing values, we re-trained all mortality-based models using the remaining 1,461 proteins The coefficients for these models are available in Supp. Table 3.

The Filbin et al. (2021), Dammer et al. (2022) and Damsky et al. (2022) datasets used the same Olink panel to measure 1,463 proteins (as opposed to the panel of 2,923 proteins that was measured in the samples from the initial visit in UK Biobank). We therefore also used these reduced models trained in UK Biobank to validate our findings in these datasets. The 1^st^-generation aging models trained directly on CSF and plasma from the Dammer et al. (2022) dataset were only trained on the 1,054 proteins that were reliably detected in CSF (with *n* = 147 and 37 individuals for the training and test datasets, respectively).

For the longitudinal analysis, rates of aging were calculated for the 1,006 participants with Olink proteomics data available for the first, third, and fourth visit, based on the following mixed-effects model:

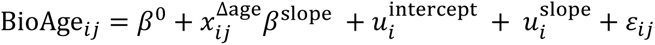

Herein, BioAge_*ij*_ denotes the predicted relative log(mortality hazard) as calculated by the mortality-based aging models for the *i*th participant on the *j*th visit, *β*^0^ is a fixed intercept, 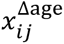 is the difference in chronological age between the *j*th visit and the first visit for the *i*th participant. The fixed effect *β*^slope^ is the overall change in predicted relative log(mortality hazard) per year of chronological age. 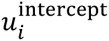 is a random effect that allows for participant-specific intercepts, 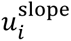 is a random effect that allows for participant-specific changes in predicted relative log(mortality hazard) per year of chronological age. *ɛ*_*ij*_ is a random error term. All random effects and the random error term are assumed to follow normal distributions. Mixed-effects models were implemented with the lmer function from lme4 R package. The longitudinal slope for each individual *i* was calculated as 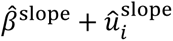 (with hats indicating the estimators for the corresponding model parameters).

Hazard ratios from the Oh et al. (2023) study for the LonGenity cohort were obtained from their Supplementary Table 12. Hazard ratios for the Lu et al. (2019) study were obtained from their Supplementary Table 5. They were standardized by dividing the regression coefficient and its confidence intervals on the log-scale by the standard deviation of their age gaps for the LonGenity cohort (Supplementary Table 8 from Oh et al. (2023)) and the standard deviation of their AgeAccelGrim phenotypes (Supplementary Table 9 from Lu et al. (2019)), respectively, to calculate standardized regression coefficients. Standardized hazard ratios and their confidence intervals were then calculated by taking the exponential function of standardized effect sizes.

### Statistical Analyses

The effects of the individual plasma proteins on chronological age after correction for sex were assessed using ordinary linear regression. More specifically, for each protein *p*, we constructed the following model:

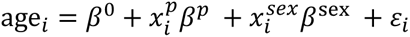

Herein, age_*i*_ is the chronological age of the *i*th individual, *β*^0^ is a fixed intercept, 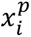 is the median-centered Olink Normalized Protein Expression (NPX) value for protein *p* and participant *i*, as provided by UK Biobank Record Table 1072. *β*^*p*^ is the effect of protein 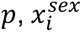 is a dummy variable equal to 1 if participant *i* is male, and 0 if participant *i* is female. *β*^sex^ is the effect of male sex, and *ɛ*_*i*_ is a random error term.

The effects of the individual plasma proteins on mortality were assessed in a similar way with Cox proportional hazards models with the time difference between death and the first visit as an outcome for the deceased subjects, and the time difference between the latest recorded death and the first visit as the outcome for the surviving subjects. Here, we included main effects for chronological age and sex, and the interaction between chronological age and sex as covariates in the model.

Gene set enrichment analysis was performed using the adaptive multilevel splitting Monte Carlo approach implemented in the fgseaMultilevel function from the fgsea R package v 1.20.0. Gene sets were obtained from MSigDB and included gene ontology: biological process (GObp), KEGG, Reactome, and Hallmarks.

The gene expression signatures of chronological aging in mouse and human blood, human tissues (multi-tissue), mouse and rat tissues (multi-tissue), and mouse, rat and human tissues (multi-tissue) were obtained from Tyshkovskiy et al. (2023). Correlation analysis was performed at the level of individual genes and proteins using slope estimates and at the level of enriched pathways using normalized enrichment scores (NES) derived from GSEA.

When only comparing two groups without covariates, we used the default two-sided two-sample t-test with Welch correction for unequal variances, as implemented in the R stats package. If covariates were included, we used ordinary linear regression. For time-to-event analyses, we used Cox proportional hazards models.

### Multiple testing corrections

We controlled for multiple testing wherever appropriate. When performing a small number of tests simultaneously (<10) within the same model framework, we controlled the family-wise error rate at 5% using the default single-step method, as implemented in the glht R package. When performing a small number of tests simultaneously (<10) outside of the same model framework, we controlled the family-wise error rate at 5% using the Hommel procedure. For larger numbers of comparisons (≥10), we controlled the false discovery rate (FDR) at 5% with the Benjamini-Hochberg FDR method.

### Software

Aging models were trained using the scikit-learn library using Python 3.10 on Jupyter Lab Version 3.2.1. All other analyses were performed in RStudio Pro 2022.12.0 Build 353.pro20 “Elsbeth Geranium” running R version 4.1.2 (2021-11-01) on a 64-bit CentOS Linux 7 server. When needed, manual editing of figures was done in Inkscape v. 1.3.2. Some color palettes used in this publication were obtained from or inspired by the MetBrewer R package.

## Acknowledgements

This research has been conducted using the UK Biobank Resource under application number 21988. We would like to thank Dr. Peter Fedichev and Dr. Timothy Pyrkov for providing us with estimates of Phenotypic Age and Locomotor Age.

## Author contributions

L.J.E.G. performed analyses, made the figures and wrote the manuscript. A.E. formatted the Anatomical Therapeutic Chemical codes and mapped them to UK Biobank medications. A.T. performed gene set enrichment analyses and correlation analyses of existing transcriptomic data. M.A.A. performed all comparisons of our study with the Oh et al. (2023) study and GrimAge. K.Y. and M.M. performed validation analyses. V.N.G. conceived the study, supervised the work, provided funding and wrote the manuscript. All authors approved the manuscript and gave critical comments.

## Data availability

The sensitive personal-level UK Biobank data used in this research cannot be made not openly available. However, all *bone fide* researchers who wish to conduct health-related research can [apply for access](https://www.ukbiobank.ac.uk/enable-your-research/apply-for-access) to UK Biobank through UK Biobank’s access management system. Filbin et al. (2021) made their data openly available at Mendeley Data (DOI: 10.17632/nf853r8xsj.2). Dammer et al. (2022) made their data openly available through the Synapse collaborative research platform at https://www.synapse.org/#!Synapse:syn30549757/files/. The Damsky et al. (2022) data is available from the Gene Expression Omnibus (GEO) under identifier GSE169148. The proteomics data from the MESA study can be obtained from dbGaP under identifier phs001416.v3.p1. The coefficients from our conventional and organ-specific models are available in the supplementary tables.

## Code availability

All analyses have been carried out using freely available software packages in python and R. All code to reproduce the analyses in this manuscript will be made available at www.github.com/ludgergoeminne/organAging.

