## Supplementary material for "Plasma-based organ-specific aging and mortality models unveil diseases as accelerated aging of organismal systems": Supp. Fig.

### Supplementary figures

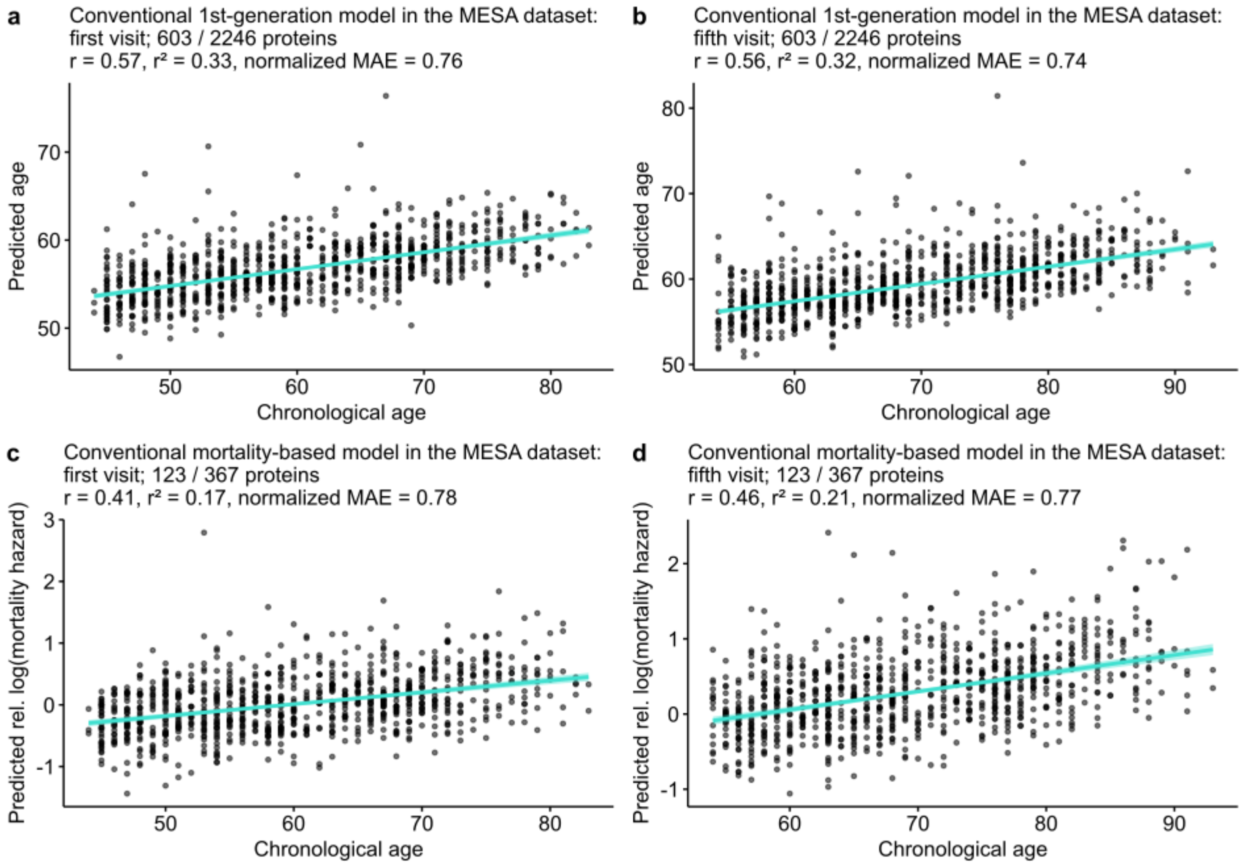


**Supplementary Figure 1. The conventional 1st-generation model and the conventional mortality-based model predict chronological age in the SomaScan proteomics from the external MESA study (Bild et al., 2002), even if not all proteins are present in the dataset. a – b.** Biological age as predicted by our 1st-generation proteome aging model has a strong positive correlation with chronological age in the MESA dataset, both in the SomaScan proteomics collected at the first (a) and the fifth visits (b) (*n* = 921 participants each). **c – d.** Biological age as predicted by our mortality-based proteome aging model has a strong positive correlation with chronological age in the MESA dataset, both in the SomaScan proteomics collected at the first (a) and the fifth visits (b) (*n* = 921 participants each). In all panels, the number of proteins from the aging models with non-zero coefficients used in the SomaScan dataset is shown as a fraction of the total number of proteins with non-zero coefficients for the aging models. r: correlation coefficient, $r^2$: coefficient of determination, normalized MAE: mean absolute error of the normalized residuals, magenta: women, blue: men. Robust regression lines with 95% confidence bands (shaded area) are added.

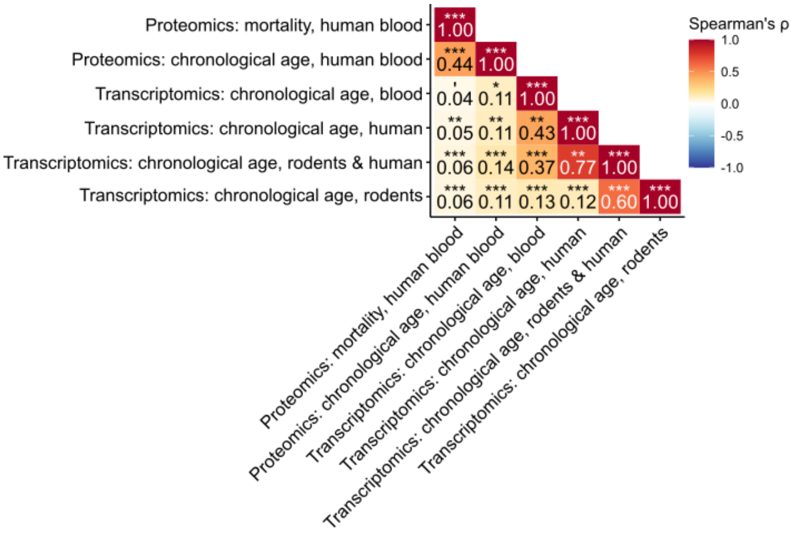


**Supplementary Figure 2. Correlation of proteomic and transcriptomic signatures/biomarkers of chronological age and mortality.** Heatmap showing the correlation coefficients (r) at the protein- or gene-level between the protein-level analysis for chronological age, the protein-level analysis for mortality, and several transcriptomic associations with chronological age derived from Tyshkovskiy et al. (2023).

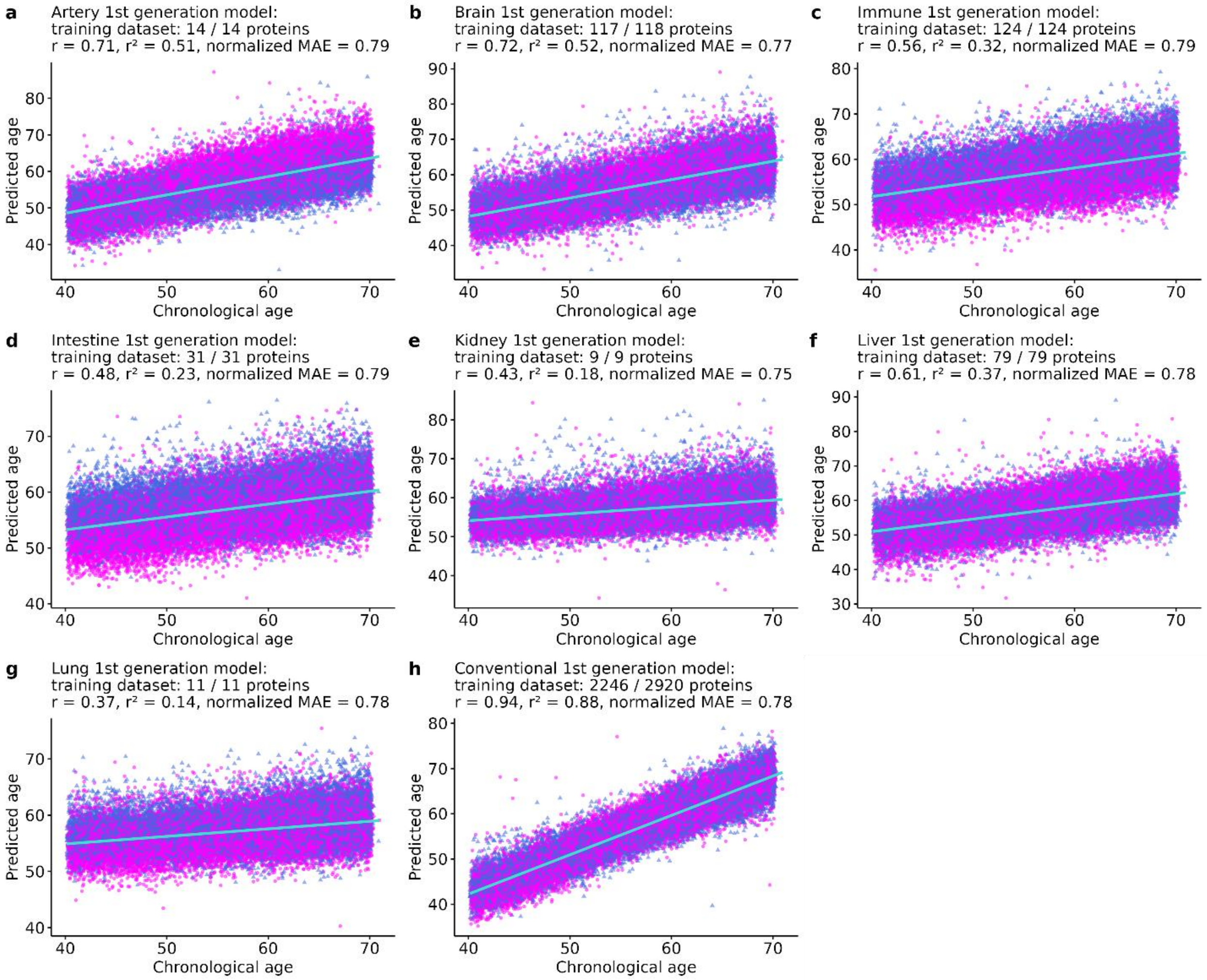


**Supplementary Figure 3. Predicted biological age correlates positively with chronological age in the training dataset (*n* = 42,412 UK Biobank participants) for all organ-specific 1$^{st}$-generation aging models.** r: correlation coefficient, r²: coefficient of determination, normalized MAE: mean absolute error of the normalized residuals, magenta: women, blue: men. Robust regression lines with 95% confidence bands (shaded area) are added.

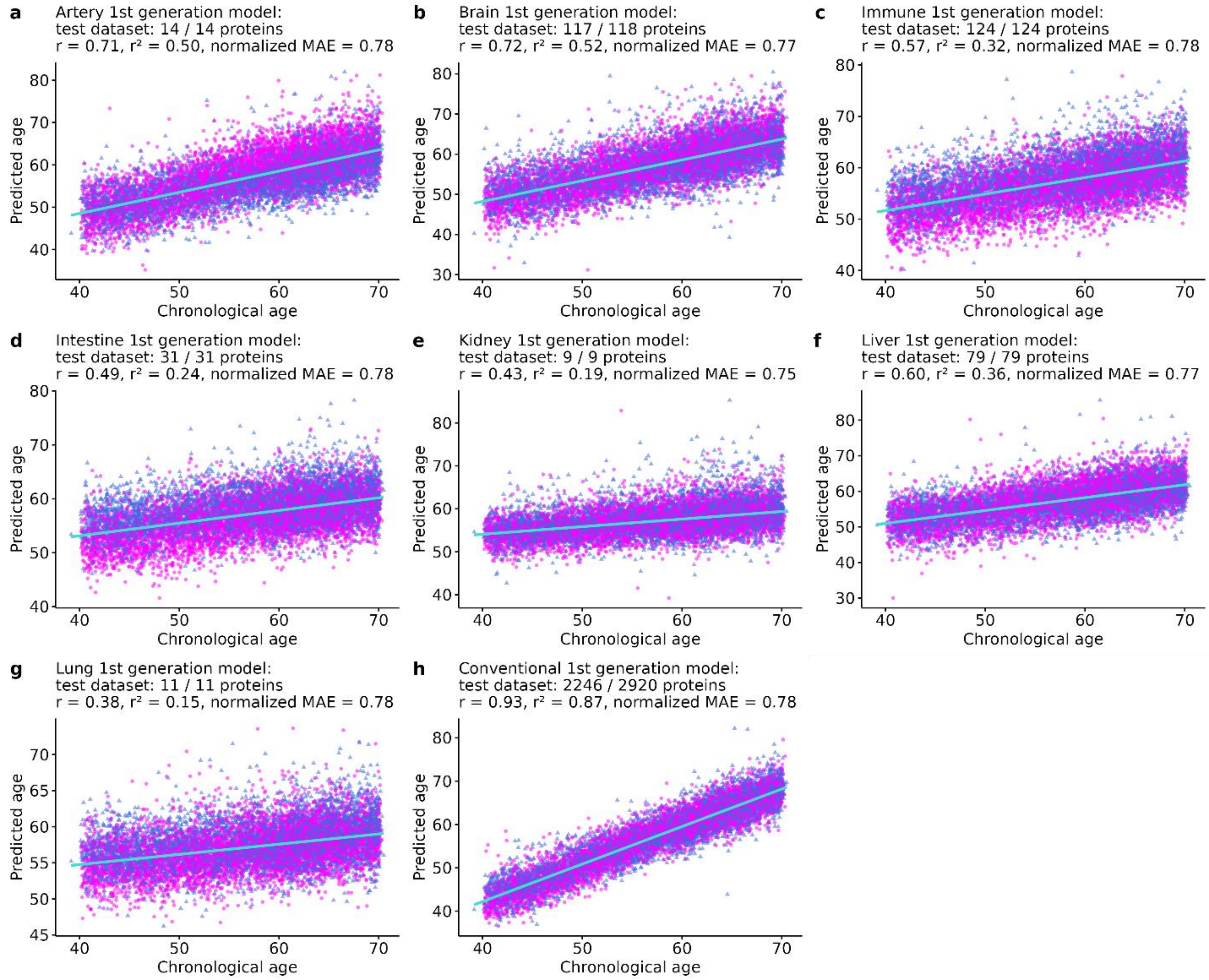


**Supplementary Figure 4. Predicted biological age correlates positively with chronological age in the test dataset (*n* = 10,603 UK Biobank participants) for all organ-specific 1st-generation aging models.** r: correlation coefficient, $r^2$: coefficient of determination, normalized MAE: mean absolute error of the normalized residuals, magenta: women, blue: men. Robust regression lines with 95% confidence bands (shaded area) are added.

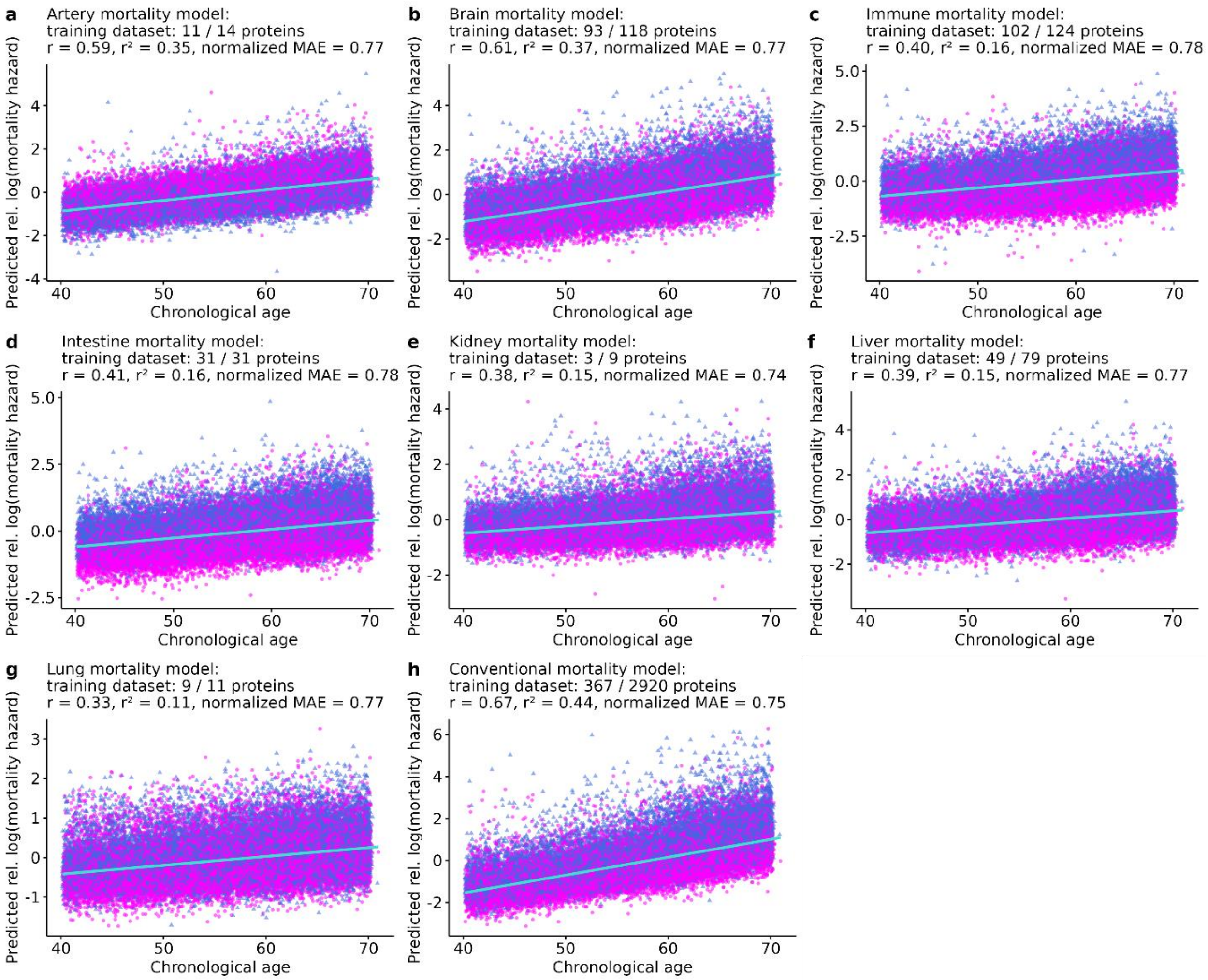


**Supplementary Figure 5. Predicted relative log(hazard) of mortality correlates positively with chronological age in the training dataset (*n* = 42,412 UK Biobank participants) for all mortality-based aging models.** r: correlation coefficient, $r^2$: coefficient of determination, normalized MAE: mean absolute error of the normalized residuals, magenta: women, blue: men. Robust regression lines with 95% confidence bands (shaded area) are added.

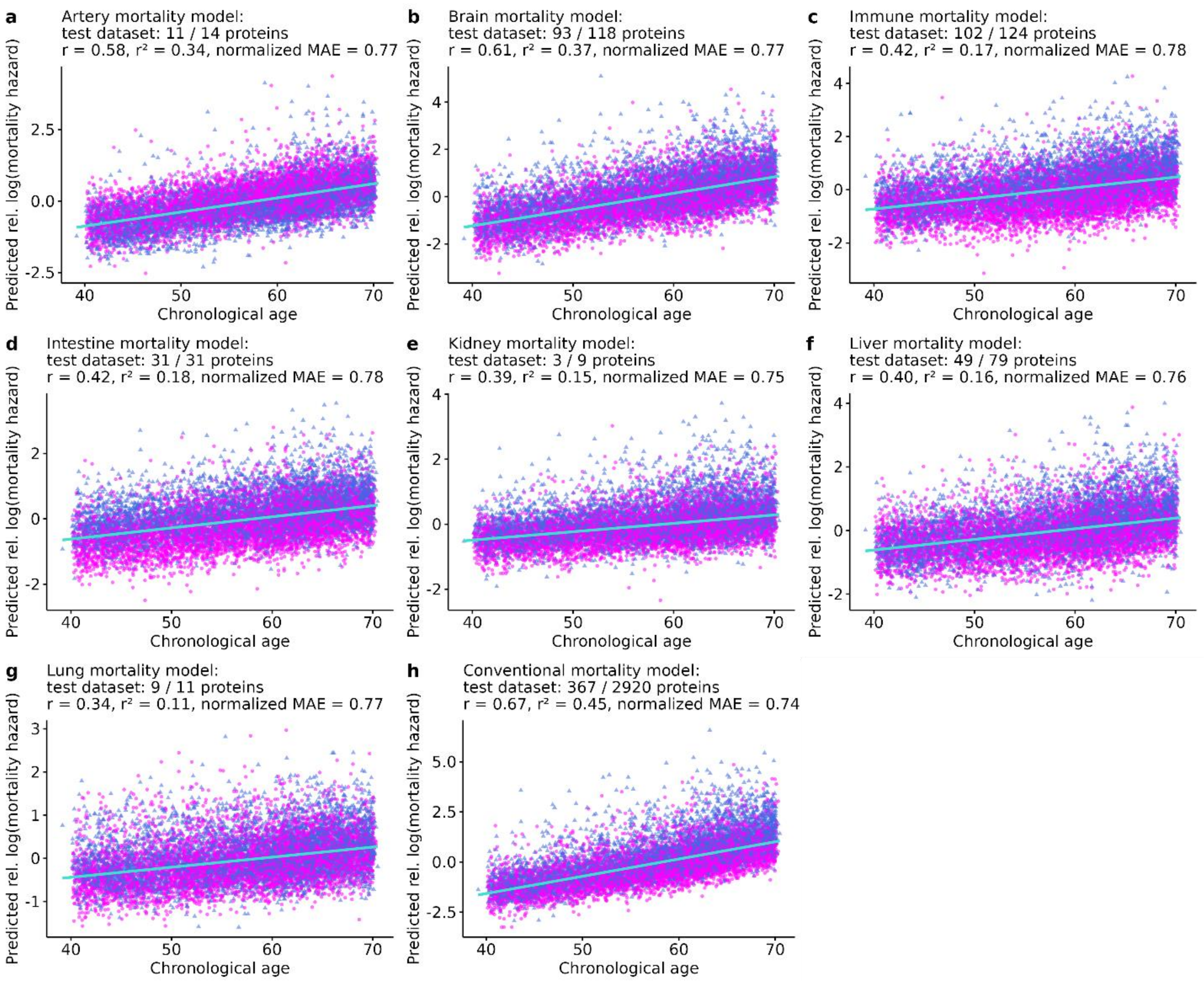


**Supplementary Figure 6. Predicted relative log(hazard) of mortality correlates positively with chronological age in the test dataset (*n* = 10,603 UK Biobank participants) for all mortality-based aging models.** r: correlation coefficient, $r^2$: coefficient of determination, normalized MAE: mean absolute error of the normalized residuals, magenta: women, blue: men. Robust regression lines with 95% confidence bands (shaded area) are added.

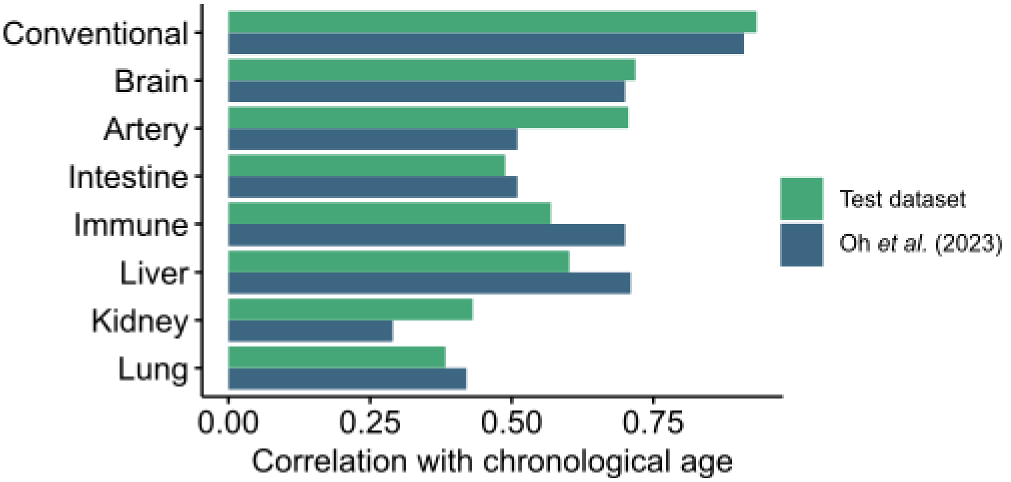


**Supplementary Figure 7. Organ-specific aging models predict chronological age.** Bar plots showing the correlation coefficients (r) with chronological age for the predicted ages by the conventional and organ-specific 1st-generation aging models in the test dataset (*n* = 10,603 UK Biobank participants), and the correlation coefficients with chronological age reported by Oh et al. (2023).

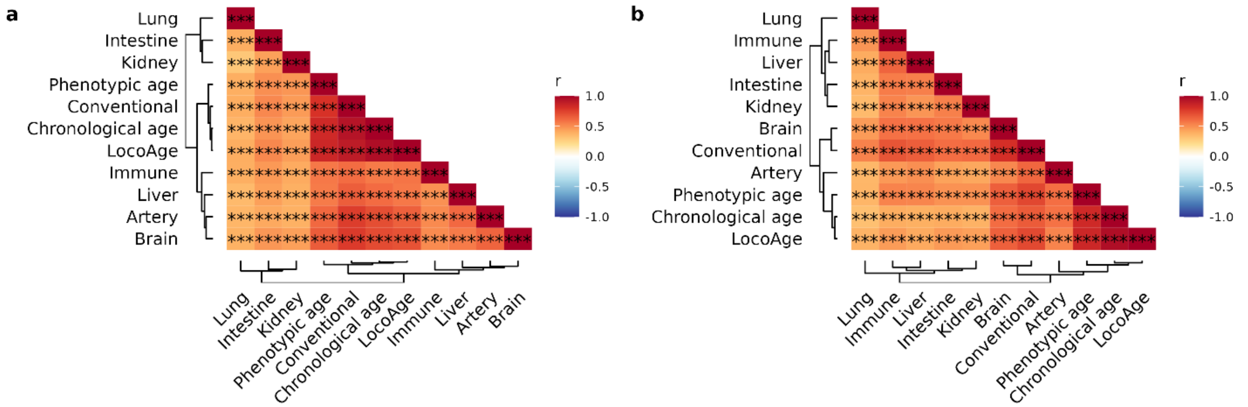


**Supplementary Figure 8. Organ-specific aging models correlate with the conventional model, chronological age, phenotypic age and locomotor age. a.** Heatmap showing the correlation coefficients (r) for the predicted ages by the organ-specific 1st-generation aging models, the conventional 1st-generation aging model, and chronological age (*n* = 53,015 UK Biobank participants), as well as phenotypic age (*n* = 44,901 UK Biobank participants) and LocoAge (*n* = 10,428 UK Biobank participants). **b.** Heatmap showing the correlation coefficients (r) for the predicted relative log(hazards) of mortality by the mortality-based organ-specific aging models, the conventional mortality-based aging model, and chronological age (*n* = 53,015 UK Biobank participants), as well as phenotypic age (*n* = 44,901 UK Biobank participants) and LocoAge (*n* = 10,428 UK Biobank participants).

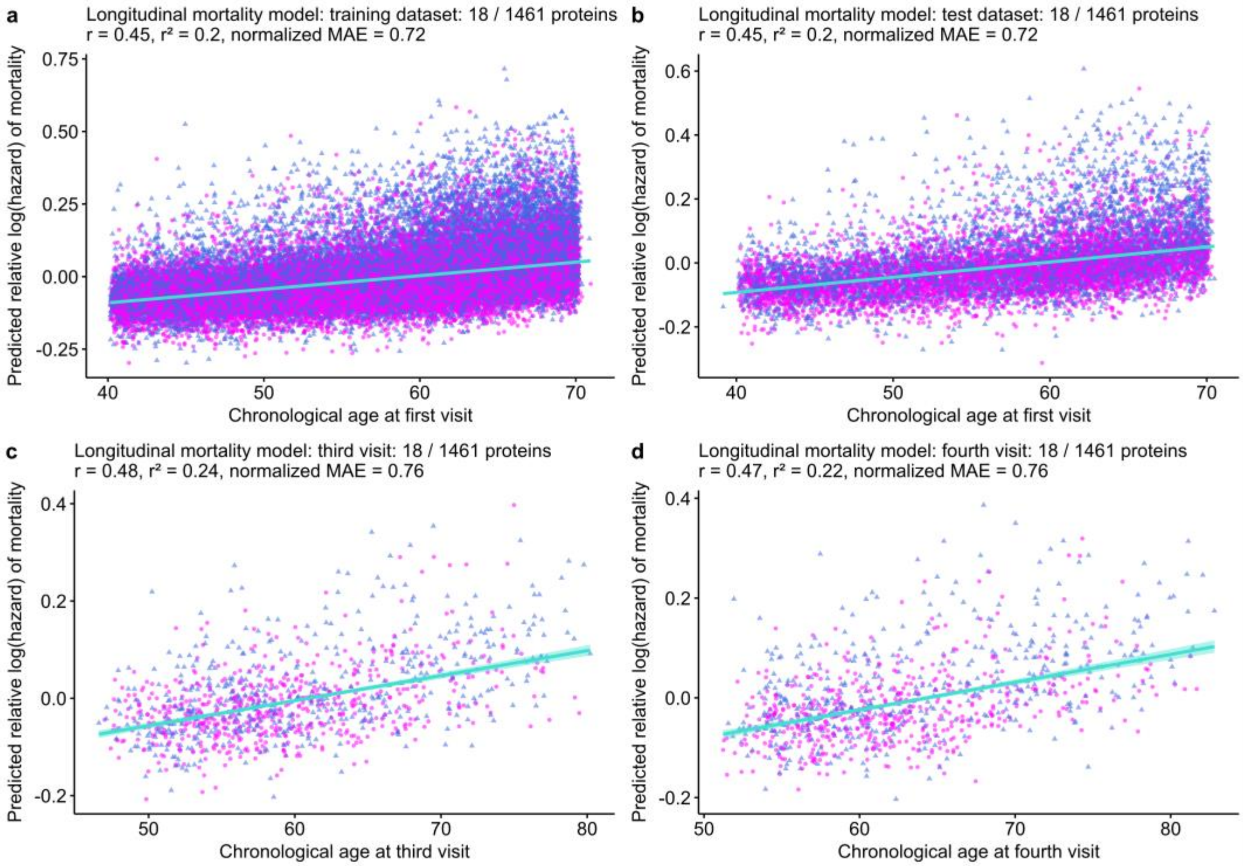


**Supplementary Figure 9. Longitudinal mortality-based models associate with chronological age. a.** Relative log(hazard) of mortality as predicted by the conventional longitudinal mortality-based aging model correlates positively with chronological age in the training dataset (*n* = 42,412 UK Biobank participants). **b.** Relative log(hazard) of mortality as predicted by the conventional longitudinal mortality-based aging model correlates positively with chronological age in the test dataset (*n* = 10,603 UK Biobank participants). **c.** Relative log(hazard) of mortality as predicted by the conventional longitudinal mortality-based aging model correlates positively with chronological age for the *n* = 1132 UK Biobank participants that have proteomics data available for their third visit. **d.** Relative log(hazard) of mortality as predicted by the conventional longitudinal mortality-based aging model correlates positively with chronological age for the *n* = 1006 UK Biobank participants that have proteomics data available for their fourth visit. r: correlation coefficient, $r^2$: coefficient of determination, normalized MAE: mean absolute error of the normalized residuals, magenta: women, blue: men. Robust regression lines with 95% confidence bands (shaded area) are added.

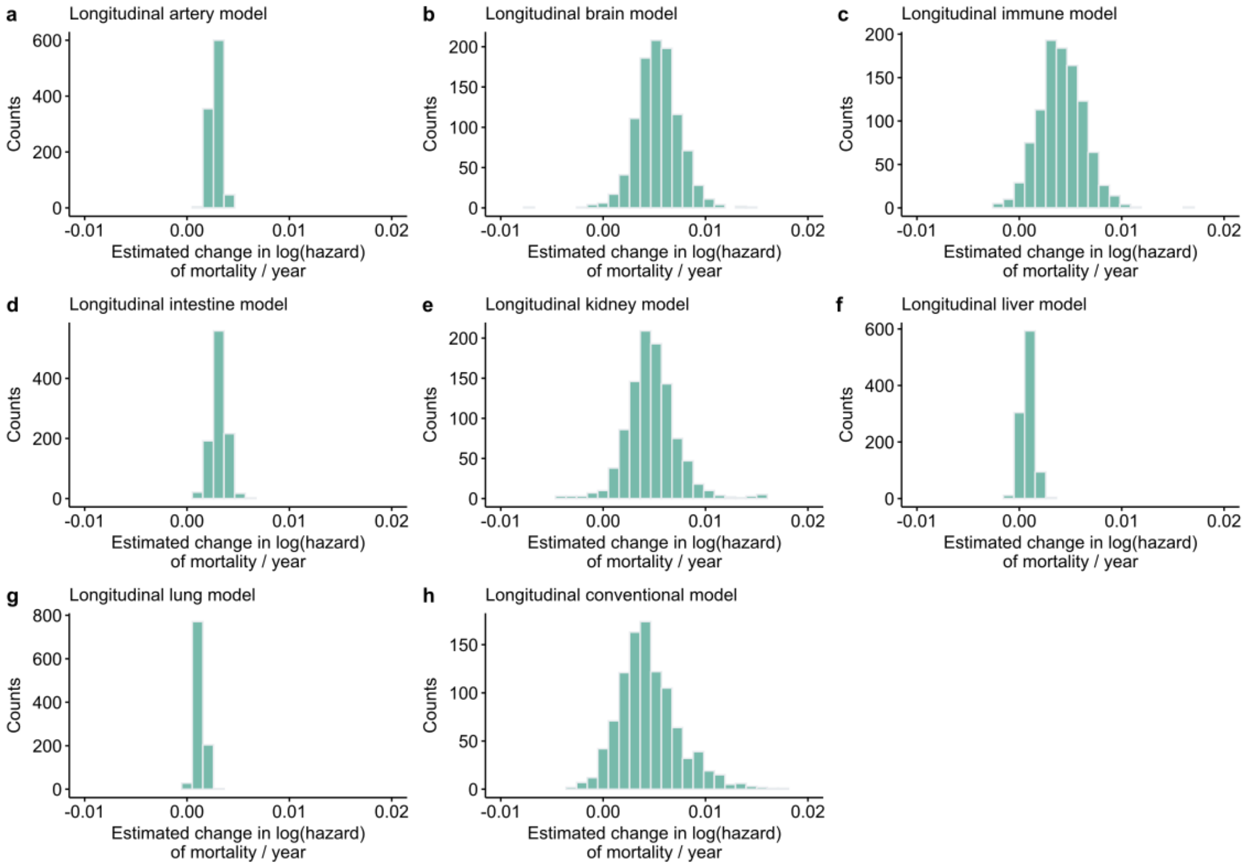


**Supplementary Figure 10. The estimated rates of aging as measured by longitudinal mortality-based aging models are mostly positive.** The histograms show the distributions of the slopes of the biological ages predicted by the conventional and organ-specific mortality-based models for the $n$ = 1,006 individuals that have proteomics data available for their first, third and fourth visits.

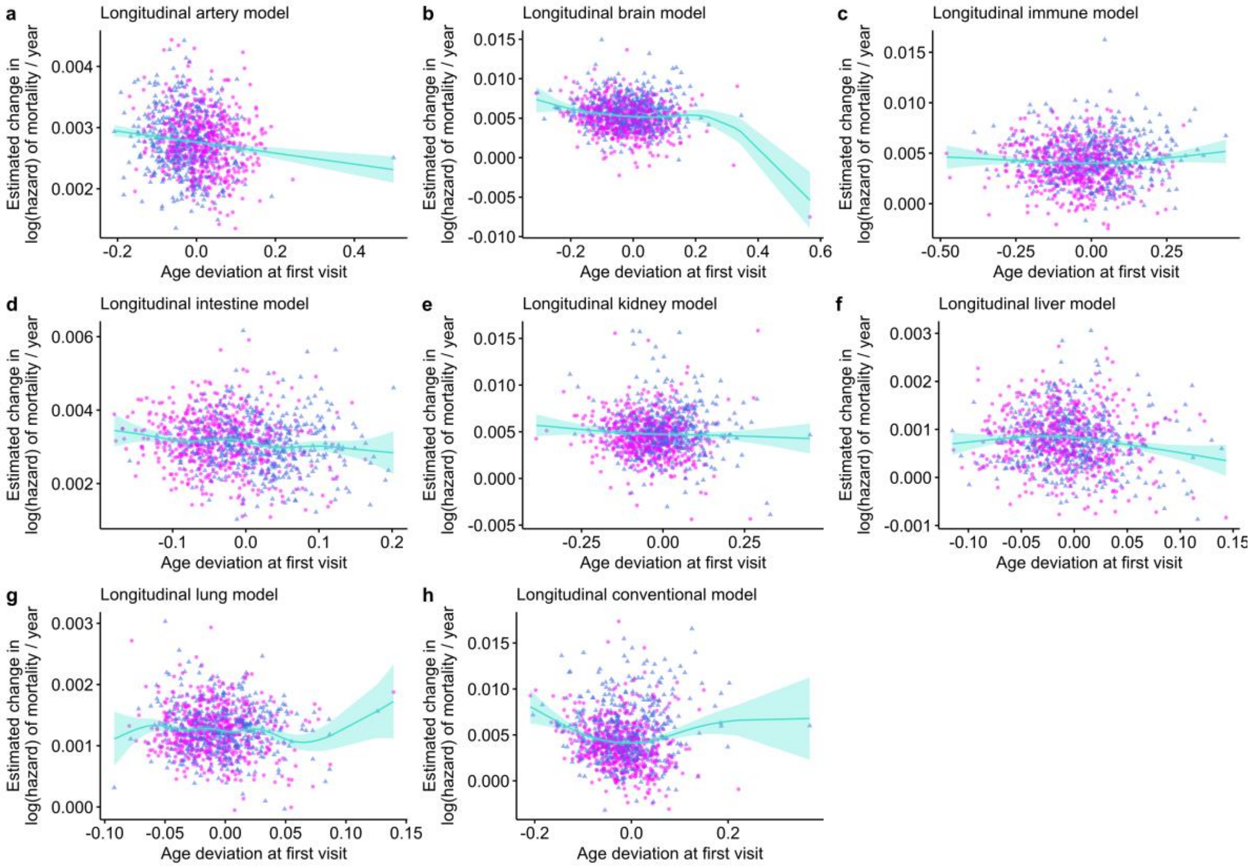


**Supplementary Figure 11. For most organ-specific mortality-based models, there is no clear association between the age deviation at first visit and the rate of aging.** Effects of age deviation at first visit on the rate of aging as measured by longitudinal mortality-based aging models.

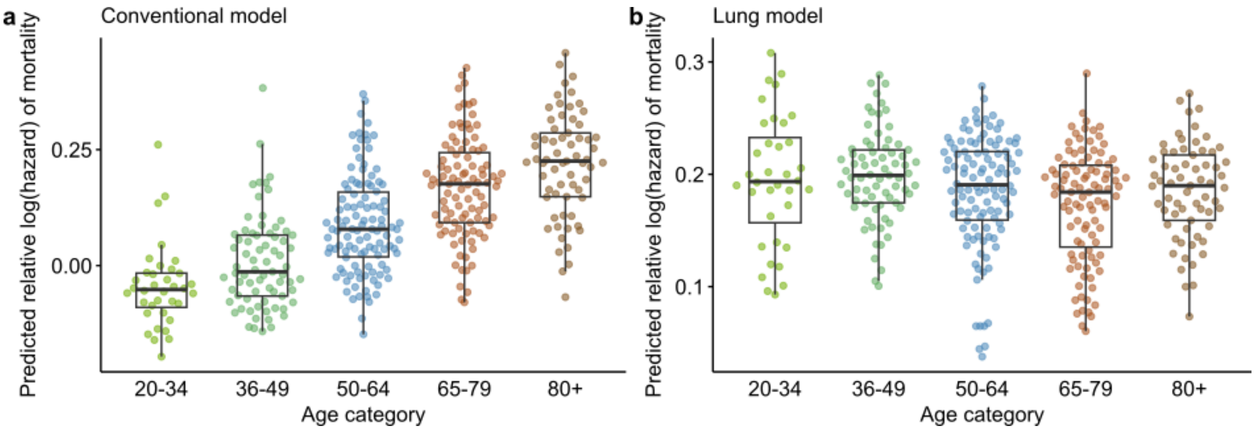


**Supplementary Figure 12. The conventional, but not the lung-specific mortality-based model associate with chronological age. a.** Data is from the Filbin et al. (2021) dataset. The mortality hazard predicted by the conventional model strongly associates with age (ordinary least squares p value < $1*10^{-16}$, r = 0.62). **b.** The mortality hazard predicted by the lung model shows a slightly negative significant age association (ordinary least squares p value = 0.01, r = 0.13).

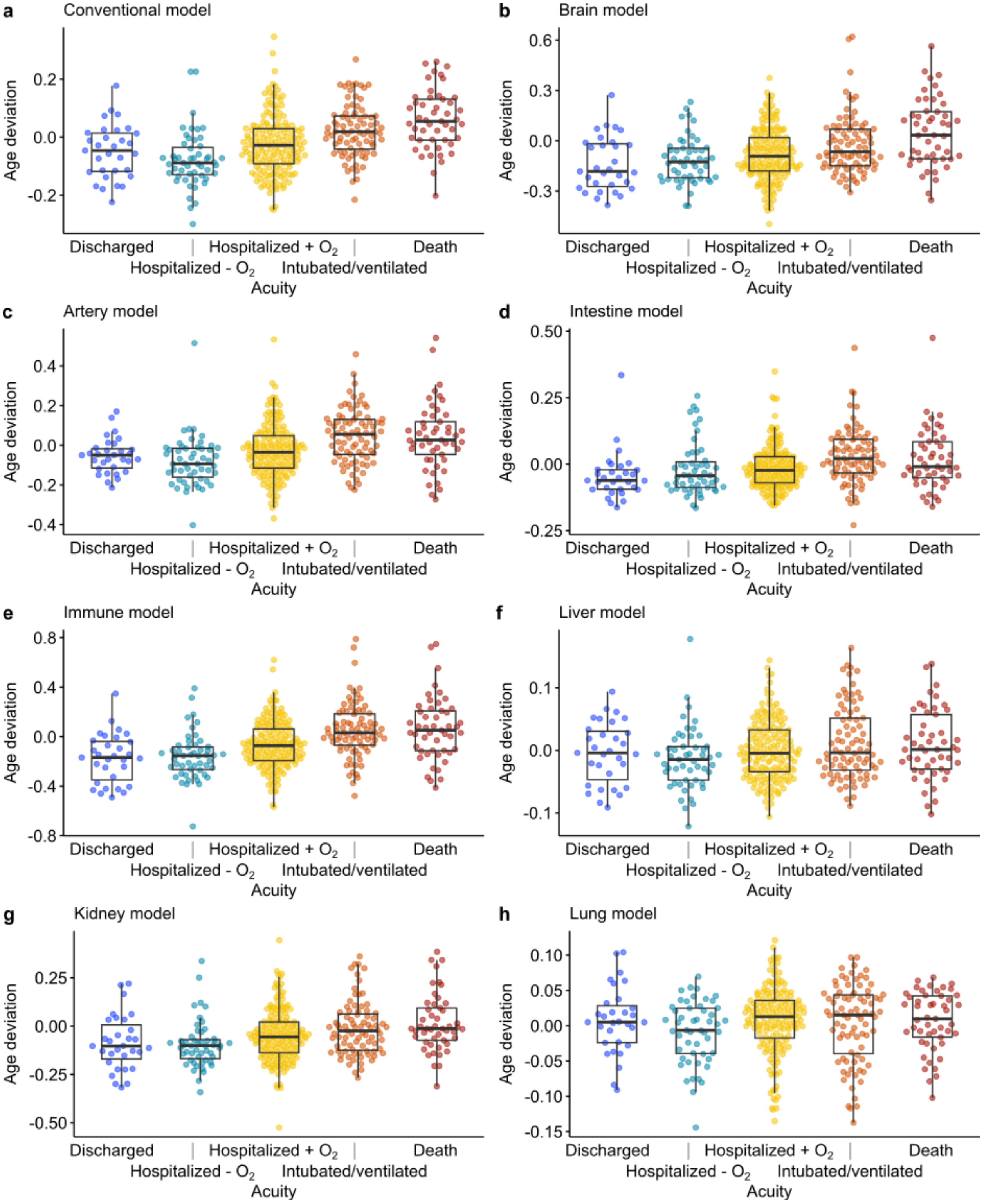


**Supplementary Figure 13. Biological age deviations as predicted by the conventional mortality-based model and all organ-specific models except lung associate with increased COVID severity.** Ordinary linear regression p-values with Hommel correction: $2*10^{-12}$ (conventional model), $7*10^{-9}$ (brain model), $9*10^{-8}$ (artery model), $1*10^{-4}$ (intestine model), $5*10^{-12}$ (immune model), 0.02 (liver model), $1*10^{-5}$ (kidney model), and 0.5 (lung model). “Discharged”: patients not hospitalized and survived to 28 days (*n* =

31), “hospitalized - $O_2$”: patients hospitalized, but no supplementary oxygen required and survived to 28 days ($n$ = 51), “hospitalized + $O_2$”: patients hospitalized, but supplementary oxygen was required and survived to 28 days ($n$ = 169), “Intubated/ventilated”: patients hospitalized and intubated and/or ventilated, and survived to 28 days ($n$ = 83). “Death”: patients died within 28 days ($n$ = 49).

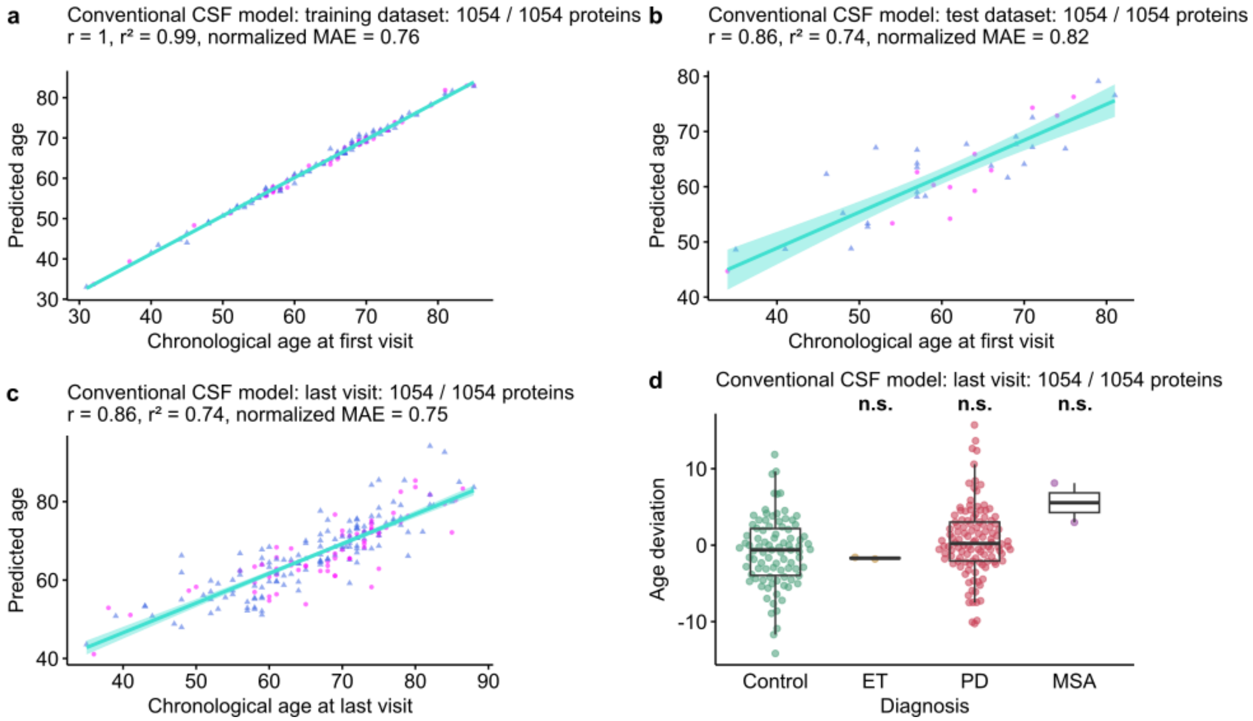


**Supplementary Figure 14. Performance of a conventional chronological aging model trained on the CSF proteome.** Data is from the Dammer et al. (2022) dataset. **a – b.** Biological ages predicted by the conventional aging model trained on CSF data correlate positively with chronological age in the CSF training and test samples ($n$ = 147 and 37 individuals for the training and test datasets, respectively). The number of proteins with non-zero coefficients is shown as a fraction of the total number of proteins on which the models are trained. r: correlation coefficient, $r^2$: coefficient of determination, normalized MAE: mean absolute error of the normalized residuals, magenta: women, blue: men. Robust regression lines with 95% confidence bands (shaded area) are added. **c.** Biological ages predicted by the conventional aging model trained on CSF data correlate positively with chronological age in the CSF samples taken at the last visit ($n$ = 212 individuals). r: correlation coefficient, $r^2$: coefficient of determination, normalized MAE: mean absolute error of the normalized residuals, magenta: women, blue: men. Robust regression lines with 95% confidence bands (shaded area) are added. **d.** Biological ages predicted by the conventional aging model trained on CSF data do not differ significantly in age deviation in the essential tremor (ET) group ($n$ = 2 individuals), the Parkinson’s disease (PD) group ($n$ = 118 individuals), and the multiple systems atrophy (MSA) group ($n$ = 2 individuals), as compared to the control group ($n$ = 90 individuals) (single-step-adjusted p = 1, 0.07, and 0.1, respectively).

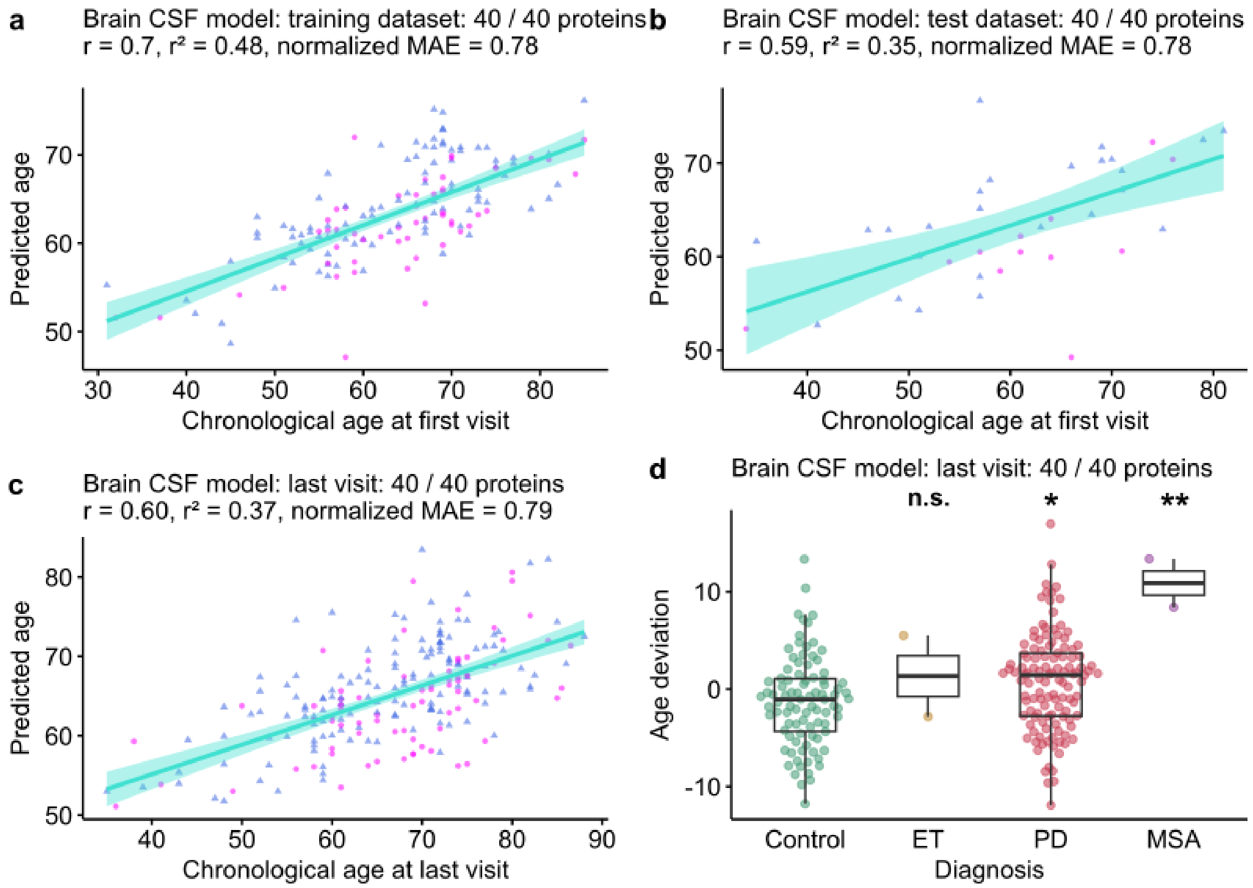


**Supplementary Figure 15. Performance of a brain-specific chronological aging model trained on the CSF proteome.** Data is from the Dammer et al. (2022) dataset. **a – b.** Biological ages predicted by the brain-specific aging model trained on CSF data correlate positively with chronological age in the CSF training and test samples (*n* = 147 and 37 individuals for the training and test datasets, respectively). The number of proteins with non-zero coefficients is shown as a fraction of the total number of proteins on which the models are trained. r: correlation coefficient, $r^2$: coefficient of determination, normalized MAE: mean absolute error of the normalized residuals, magenta: women, blue: men. Robust regression lines with 95% confidence bands (shaded area) are added. **c.** Biological ages predicted by the brain-specific aging model trained on CSF data correlate positively with chronological age in the CSF samples taken at the last visit (*n* = 212 individuals). r: correlation coefficient, $r^2$: coefficient of determination, normalized MAE: mean absolute error of the normalized residuals, magenta: women, blue: men. Robust regression lines with 95% confidence bands (shaded area) are added. **d.** Biological ages predicted by the brain-specific aging model trained on CSF data do not differ significantly in age deviation in the essential tremor (ET) group (*n* = 2 individuals), the Parkinson's disease (PD) group (*n* = 118 individuals), and the multiple systems atrophy (MSA) group (*n* = 2 individuals), as compared to the control group (*n* = 90 individuals) (single-step-adjusted p = 0.8, 0.01, and 0.002, respectively).

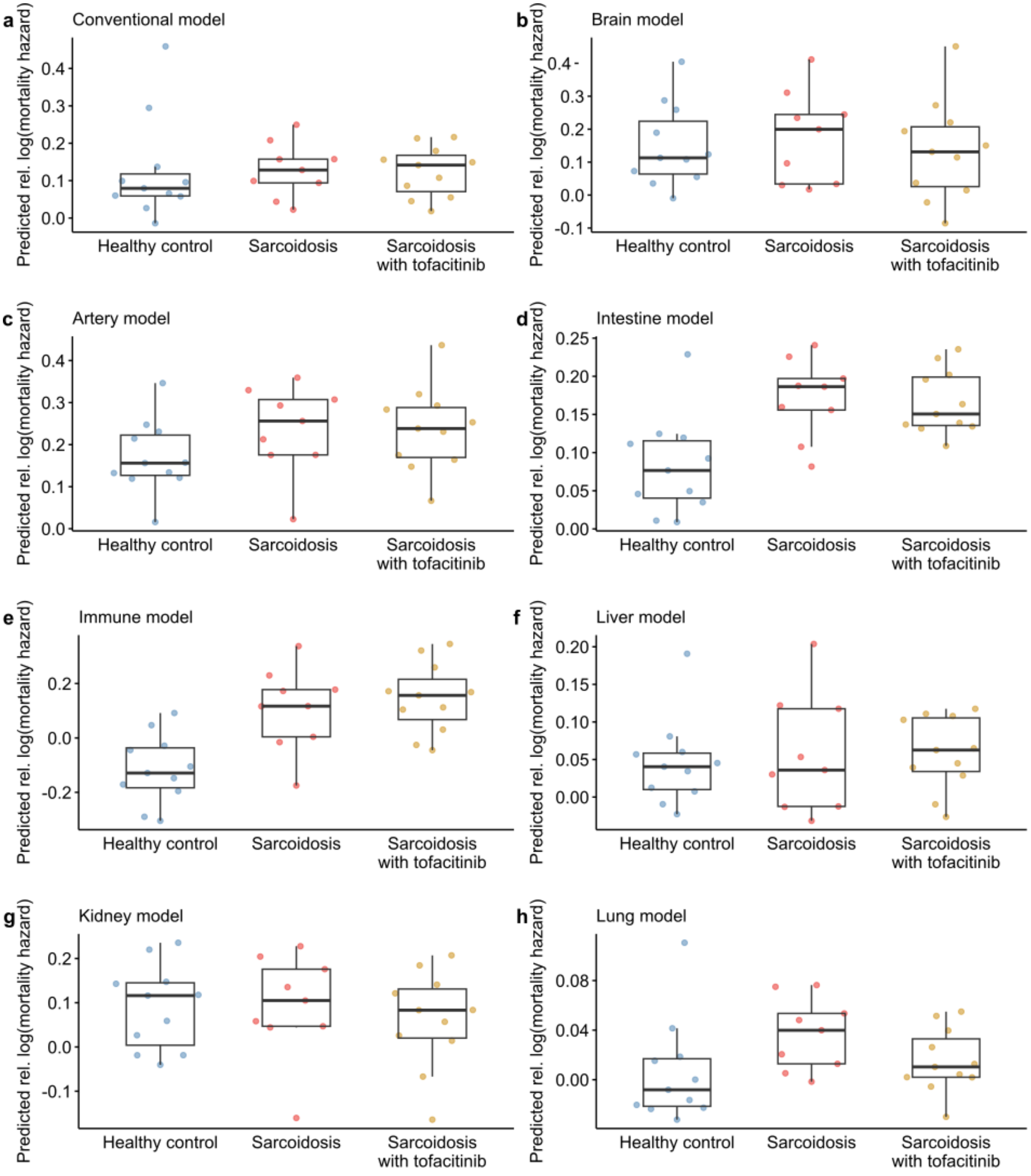


**Supplementary Figure 16. The mortality-based immune and intestine aging models associate with sarcoidosis.** Relative log(hazards) of mortality as predicted by the mortality-based conventional and organ-specific aging models based on the plasma proteomics of 11 healthy controls, 9 untreated sarcoidosis patients, and 11 sarcoidosis patients treated with tofactinib, measured with the Olink Explore 1536 platform in the dataset of Damsky et al. (2022). Ordinary linear regression p-values with Hommel correction for the comparison sarcoidosis with or without tofacitinib vs healthy controls: 0.9 (conventional model), 0.9 (brain model), 0.4 (artery model), 0.001 (intestine model), 0.001 (immune model), 0.9 (liver model), 0.9 (kidney model), and 0.6 (lung model).
